# Distinct Adaptive Immunophenotypes in duodenal mucosa but not in peripheral blood of patients with functional dyspepsia

**DOI:** 10.1101/2021.11.22.21266508

**Authors:** Grace L. Burns, Jessica Bruce, Kyra Minahan, Andrea Mathe, Thomas Fairlie, Raquel Cameron, Crystal Naudin, Prema M. Nair, Michael D. E. Potter, Mudar Zand Irani, Steven Bollipo, Robert Foster, Lay T. Gan, Ayesha Shah, Natasha Koloski, Paul S. Foster, Jay Horvat, Martin Veysey, Gerald Holtmann, Nick Powell, Marjorie M. Walker, Nicholas J. Talley, Simon Keely

**Affiliations:** College of Health, Medicine and Wellbeing, The University of Newcastle, Callaghan, NSW, Australia; Hunter Medical Research Institute, New Lambton Heights, NSW, Australia; NHMRC Centre of Research Excellence in Digestive Health, University of Newcastle, Newcastle, NSW, Australia; Translational Research Institute, Brisbane, QLD, Australia; Department of Gastroenterology, John Hunter Hospital, Newcastle, New South Wales, Australia; Department of Gastroenterology and Hepatology, Princess Alexandra Hospital, Brisbane, Queensland, Australia; Hull-York Medical School, University of Hull, Hull, United Kingdom; Imperial College London, London, United Kingdom

**Author notes:** Correspondence to: Simon Keely, Ph.D. Authors contributed equally (Nicholas J. Talley, Simon Keely).

**Keywords:** Functional dyspepsia, Functional gastrointestinal disorder, T cells, Lymphocytes, Immunology

## Abstract

**Background and aims:** Functional dyspepsia is characterised by chronic symptoms of post- prandial distress or epigastric pain not associated with defined structural pathology. Increased peripheral gut-homing T cell have been previously identified in patients. To date, it is unknown if these T cells were antigen-experienced, or if a specific immunophenotype was associated with FD. This study aimed to characterise immune populations in the blood and duodenal mucosa of FD patients that may be implicated in disease pathophysiology.

**Methods:** We identified duodenal T cell populations from 23 controls and 49 Rome III FD patients by flow cytometry. We also analysed duodenal eosinophils and T cell populations in peripheral blood from 37 controls and 49 patients and investigated if subtyping patients based on reported symptoms or co-morbidity identified specific immunophenoptypes.

**Results:** In addition to increased duodenal mucosal CD4^+^ effector cells, FD patients demonstrated a shift in the T helper cell balance compared to controls. Patients had increased duodenal mucosal Th2 populations in the effector (13.03±16.11, 19.84±15.51, *p*=0.038), central memory (23.75±18.97, 37.52±17.51, *p*=0.007) and effector memory (9.80±10.50 vs 20.53±14.15, *p*=0.001) populations. Th17 populations were also increased in the effector (31.74±24.73 vs 45.57±23.75, *p*=0.03) and effector memory (11.95±8.42 vs 18.44±15.63, *p*=0.027) subsets.

**Conclusion:** Our findings confirm the involvement of adaptive responses in the aetiopathogenesis of FD, specifically a Th2 and Th17 signature in the duodenal mucosa. The presence of effector and memory cells suggest that the microinflammation in FD is antigen driven.

## INTRODUCTION

Functional dyspepsia (FD) is a disorder of gut-brain interaction (DGBI), formally referred to as functional gastrointestinal disorders (FGIDs), presenting with chronic gastrointestinal (GI) symptoms but lacking discernible pathology or biochemical abnormalities.[1] In the absence of treatable pathology, FD is sub-classified into symptom groups according to the Rome criteria: post-prandial distress syndrome (PDS) and epigastric pain syndrome (EPS), with the aim of directing therapeutic intervention towards symptoms.[2] Subtle pathologies are apparent in FD, with duodenal eosinophilia identified in several global cohorts.[3, 4, 5, 6, 7, 8] and increased proportions of peripheral gut-homing T cells are also reported.[9] Defects in the mucosal barrier of the duodenum, such as lower transepithelial electrical resistance[6] and increased permeability [6, 10, 11] are also reported. However, it remains unclear as to whether these defects are common to specific patient subgroups.

The cause of these pathologies is unknown, but stimulation of the immune system by luminal antigens may promote a loss of homeostasis and drive the development of subtle inflammation. Increased T cell homing suggests an adaptative immune response in the dysregulation observed in FD and patients have a greater proportion of peripheral naïve/activated (CD45RA^+^CD45RO^+^) T cells,[12] while the proportion of peripheral T helper (Th) lymphocytes remains unchanged,[9, 12] suggesting that activation of effector cells is associated with symptoms. Th cells can be sub-classified into phenotypically distinct subsets,[13] and Th2 and Th17 responses have been hypothesised as dominant in FD, given the capacity of these responses to recruit eosinophils to the duodenum.[14] However to date, alterations in these subpopulations have not been shown in FD.[15] Further, no studies have demonstrated a conclusive relationship between Rome symptom subtypes and T cell subsets.

FD often overlaps with the irritable bowel syndrome (IBS),[16] and while patients with multiple DGBIs have higher symptom severity scores and lower quality of life,[16, 17] it is unclear whether overlap patients exhibit greater homeostatic imbalance than patients with FD only. As such, we hypothesised that a loss of GI homeostasis in FD patients allows for the induction of specific T cell responses that promote chronic GI symptoms. This study aimed to characterise T helper lymphocyte subsets within the blood and mucosa of FD patients to identify specific immunological subsets of this condition. In addition, we also aimed to investigate whether subtyping of patients based on Rome III symptom criteria or the presence of concomitant irritable bowel syndrome (IBS) was associated with specific profiles.

## METHODS

### Cohort recruitment

Participants aged 18-80 years were recruited through outpatient gastroenterology clinics at John Hunter, Gosford and Wyong Hospitals in New South Wales and Princess Alexandra Hospital, Brisbane, Queensland, Australia. All research was undertaken in accordance with approvals from the Hunter New England (reference 13/12/11/3.01) and Metro South Health (reference HREC/13/QPAH/690) Human Research Ethics Committees. Patients met the Rome III criteria for PDS, or EPS with or without PDS (EPS±PDS). Asymptomatic controls required endoscopy for routine care, such as for unexplained iron deficiency anaemia (IDA), positive fecal occult blood test (+FOBT), gastro-esophageal reflux disease (GERD) or dysphagia, with no organic GI disease confirmed during endoscopy.

Exclusion criteria for the study included patients with a body mass index (BMI)>40, organic GI conditions and pregnant women. A medical interview captured medication, family history and demographic information. A validated outpatient questionnaire was also completed, incorporating the modified Nepean Dyspepsia Index[18] and Rome III questions. At endoscopy, seven biopsies were collected from the second portion of the duodenum (D2) and whole blood (36mL) was collected in lithium heparin.

### Patient and public involvement

There were no funds or time allocated for patient and public involvement so we were unable to involve patients in the intial study design. We are now engaging with patients through our NHMRC Centre for Research Excellence to develop our research and dissemination strategies.

### Isolation of peripheral blood mononuclear and duodenal lamina propria mononuclear cells

Density gradient centrifugation was performed to isolate peripheral blood mononuclear cells (PBMCs) and plasma, as previously described.[19] Plasma by-product was aliquoted and stored at -80°C for future analysis. Lamina propria mononuclear cells (LPMCs) were isolated from 5 biopsies by digestion with collagenase and DNAse II, as previously described[19] within 2 hours of sample collection.

### Surface marker staining for flow cytometry

Following thawing, cells were rested at 37°C/5% CO_2_ overnight. LPMC supernatant was collected and stored at -80°C. Cells were resuspended at 5.0x10^5^ to 8.0x10^5^ cells/mL and incubated with a fixable viability dye (conjugated to AF700), and Fc block antibody. Cells were stained (4°C/30 minutes) with the antibodies outlined in **Supplementary Table 1**, sourced from BD Biosciences, Franklin Lakes, New Jersey, USA. 4% paraformaldehyde was used to fix samples and expression of surface markers was acquired on an LSRFortessa^TM^ X20 flow cytometer with FACSDiva software (BD Biosciences). Acquired data was analysed using FlowJo v.10 (BD Biosciences) and CD3^+^ lymphocyte populations identified as outlined in **Supplementary Table 2.**

### Cytometric bead array for analysis of cytokines

The levels of IL-10, TNF, IFN-γ and IL-17A in plasma and supernatants from unstimulated LPMCs were measured using the Human TH1/TH2/Th17 Cytometric Bead Array (CBA) kit (BD Biosciences), as per manufacturer’s instructions. Data was acquired on an LSRFortessa^TM^ X20 flow cytometer with FACSDiva software (BD Biosciences). Data was analysed using FCAP array^TM^ version 3.0 software (BD Biosciences). Supernatant concentrations were normalised by dividing concentration by cell number to account for differences due to proliferation or death.

### Histology

Two formalin fixed, paraffin embedded (FFPE) D2 biopsies were stained with haematoxylin and eosin (H&E).[7] Slides were digitalised using Aperio AT2 (Leica Biosystems, Wetzlar, Germany). Eosinophil counts were performed using Aperio ImageScope (Leica Biosystems). Five high-power fields (HPFs) within a grid of 200um2, were counted in biopsies at 40x.

HPFs were selected by scanning the section for areas of increased eosinophils and counting five areas of the highest density or the single observed area of highest density and surrounding HPFs. The mean number of eosinophils per 5HPFs were calculated.

Intraepithelial lymphocytes (IELs) were counted per 50 enterocytes across 3 villi and reported as the average per 50 enterocytes.

### Immunohistochemical staining

Immunohistochemical staining was performed by NSW Regional Biopsecimen & Research Services using the Discovery Ultra Benchmark Automated Platform (Ventana Medical Systems, Inc). Cell Conditioning-1 reagent (pH 9.0, Roche, Switzerland) was used for antigen retrieval and sections were incubated for 28minutes at 37°C with anti-CD117 primary antibody (1:600, reference 4502 Aligent Dako Technologies, California, USA). Slides were incubated with an anti-rabbit secondary HQ (Roche, Switzerland) and tertiary horseradish peroxidase HQ (Roche, Switzerland). 3,3’-Diaminobenzidine (DAB) liquid substrate System (Roche, Switzerland) was used to develop sections, counterstained with haematoxylin. Slides were digitised as described, and CD117^+^ cells were counted in 5 x HPFs within a grid of 200um2, at 40x.

### Statistical analysis

Datasets were analysed using Graphpad Prism 9.2 (Graphpad Software Inc., La Jolla, USA). Characteristics of the cohort were analysed by *t-*tests. This data is presented as mean±SD. Fisher’s exact test was used to analyse effects of confounders. These data are presented as percentage of total cohort positive for tested variable.

Grubb’s outlier test was used to exclude significant outliers. Comparisons between groups were analysed by parametric *t*-tests or one-way ANOVA with uncorrected Fisher’s LSD post-hoc test for normally distributed data (assessed by D’Agostino & Pearson test). Non-normally distributed data was analysed by non-parametric *t-*tests or Kruskal-Wallis test with Dunn’s multiple comparisons post-hoc test as appropriate. This data is presented as mean±SD and *p*<0.05 was considered significant.

## RESULTS

### Cohort characteristics

#### Systemic T cell populations

PBMCs were collected from 37 controls and 61 patients (**Table 1**). Twenty patients had symptoms consistent with PDS, 6 had EPS and 35 had EPS/PDS overlap (n=41 EPS±PDS). Twelve controls were referred to endoscopy for IDA, 4 with dysphagia, 1 with reflux symptoms and 20 were undergoing FOBT screening. No patients or controls had evidence of gastrointestinal disease upon endoscopy or by histological assessment. FD patients were younger than controls (56.49±12.52 vs 48.57±16.38, *p*=0.014) but there were no differences in sex or BMI. Proton pump inhibitor (PPI) usage was higher in FD patients compared to controls (10.81% vs 69.81%, *p*<0.000) and the prevalence of co-morbid Rome III IBS was significantly lower in controls (5.41% vs 45.90%, *p*<0.000). Where available, duodenal eosinophils were counted from this cohort.

#### Duodenal biopsies

Duodenal biopsies were collected from 23 controls and 49 FD patients during upper endoscopy (**Table 2**). Twenty-four patients had PDS, 6 had EPS and 20 had EPS/PDS overlap (n=26 EPS±PDS). Indications for endoscopy among controls included IDA (n=5), dysphagia (n=3), reflux (n=5) and +FOBT (n=10). There were more females in the FD cohort (47.83% vs 77.55%, *p*=0.03) and patients had a lower BMI (28.99±5.325 vs 25.92±4.136, *p*=0.007). The prevalence of co-morbid IBS was increased in FD (4.35% vs 30.00%, *p*=0.015).

### FD patients have increased duodenal eosinophil counts

Examination of duodenal sections revealed no overt pathology comparing controls and FD subjects by H&E staining (**Figure 1A**) or CD117 (**Figure 1B**). However, duodenal eosinophil numbers were significantly increased in biopsies from FD patients (n=19, 16.95±8.05 vs n=67, 22.93±12.25, *p*=0.048) compared to control subjects (**Figure 1C).** No difference in eosinophil counts were observed when patients were stratified into subtypes or in the numbers of IELs (**Figure 1D**) or mast cells (**Figure 1E**) between control and FD. These findings confirm that our cohort has the primary known histological feature of FD, duodenal eosinophilia.

**Figure 1:**
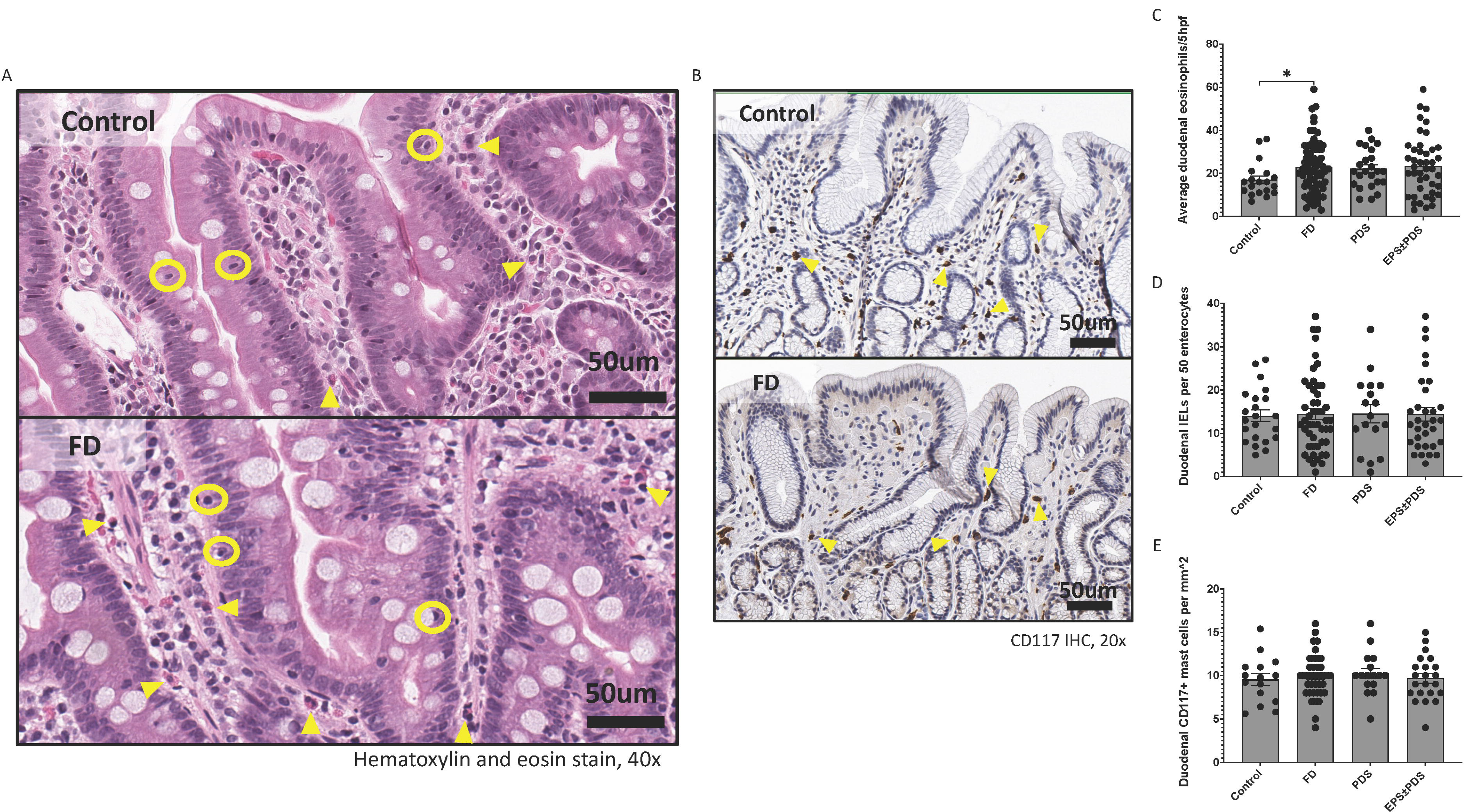
Immune hallmarks of functional dyspepsia (FD) in this cohort. Biopsies were collected from the second portion of the duodenum (D2) of FD patients and controls and stained with (A) haematoxylin and eosin (40x magnification, arrows indicate example eosinophils, circles indicate example intra-epithelial lymphocytes (IELs), scale bar=50μm). (B) CD117 immunohistochemical staining of CD117^+^ mast cells was also performed (20x magnification, arrows indicate example CD117^+^ cells, scale bar=50μm). (C) Average duodenal eosinophil counts per 5 high powered fields in control, FD, PDS and EPS±PDS patients and (D) duodenal IELs were counted across groups using H&E-stained sections. (E) Mast cells were counted based on CD117^+^ positively stained cells (brown) in all groups. n=15-22 for controls, n=38-67 for FD. Data presented as mean±SEM. Statistical analysis for control vs FD, (D, E) parametric and (C) non-parametric t test. For control vs PDS vs EPS±PDS, (E) parametric and (C, D) non-parametric one-way ANOVA. *p<0.05.

### CD4^+^ effector T cells are increased in the duodenum of FD patients

Given the duodenum is implicated as a site for symptom generation in FD, we investigated if altered lymphocyte profiles could be detected in the mucosa. There were no differences in the proportions of CD3^+^, CD4^+^ or CD8^+^ cells when FD patients were compared to controls, although EPS±PDS patients had a greater proportion of duodenal CD3^+^ lymphocytes compared to PDS (46.19±14.89 vs 37.28±12.76, *p*=0.024) (**Supplementary Figure 1**). We investigated the proportions of CD4^+^ naïve, effector, central memory (CM) and effector memory (EM) subsets based on the expression of CD45RA, CD45RO and CCR7 (**Figure 2A**) and the proportions of these subsets within the CD8^+^ population (**Figure 2B**). When the balance of each effector and memory population were considered (**Figure 2C**), there was expansion of the duodenal CD4^+^ effector compartment in FD patients. This was confirmed statistically, with increased proportions of CD4^+^ effector cells in FD (2.06±2.17 vs 3.94±2.93, *p=*0.006), attributable to EPS±PDS (3.13±2.34, *p*=0.001 vs con) (**Figure 2D**). The proportion of CD8^+^ effector cells was decreased in PDS compared to controls (2.15±3.95 vs 0.53±0.34, *p*=0.009 (**Figure 2E**), and these patients also had decreased CD4^+^ CM cells compared to controls (3.35±2.68 vs 1.93±1.78, *p*=0.013) (**Figure 2F**). EPS±PDS patients had significantly higher CD8^+^ CM cells compared to PDS (3.72±2.15 vs 2.33±2.18, *p*=0.025) (**Figure 2G**), however there were no differences in CD4^+^ or CD8^+^ naïve or EM cells in FD compared to controls (**Supplementary Figure 2**).

**Figure 2:**
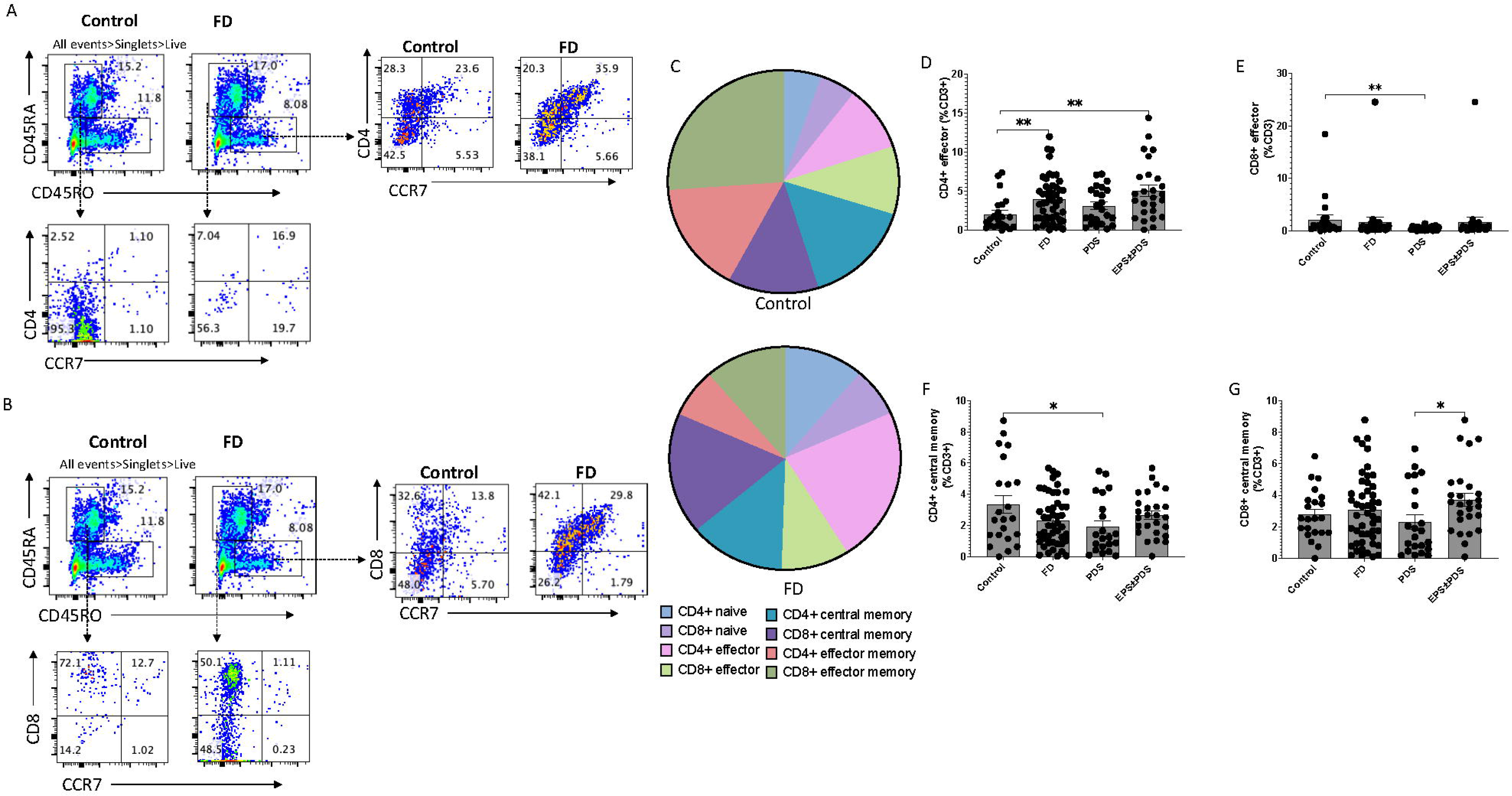
The duodenal effector and memory T cell balance in functional dyspepsia patients compared to controls. Lamina propria mononuclear cells were isolated from duodenal biopsies and phenotyped using surface marker staining and flow cytometry. Naïve (CD45RA^+^ CCR7^+^), effector (CD45RA^+^ CCR7^-^), central memory (CD45RO^+^ CCR7^+^) and effector memory (CD45RO^+^ CCR7^-^) T cells were identified in the (A) CD4^+^ and (B) CD8^+^ populations. (C) The balance of each effector and memory population was then represented within the total control and FD cohorts before the (D) CD4^+^ and (E) CD8^+^ effector populations were compared between controls and FD, as well as within the FD subtypes, PDS and EPS±PDS. The (F) CD4^+^ central memory and (G) CD8^+^ central memory populations were also investigated within these groups. n=23 controls, n=49 FD. Data presented as mean±SEM. Statistical analysis for control vs FD, (F,G) parametric t test, (D,E) non-parametric t test. For control vs PDS vs EPS±PDS, (F,G) parametric one-way ANOVA (D,E) non-parametric one-way ANOVA. *p<0.05, **p<0.01.

### Peripheral effector and memory T cell profiles do not reflect duodenal profiles

Given studies have demonstrated gut-homing cells are increased in the periphery of FD patients, we sought to characterise the circulating CD4^+^ cell populations using flow cytometry. There was no difference in CD4^+^ 4^+^ 7^+^CCR9^+^ populations between FD and controls, however EPS±PDS patients had increased CD4^+^ gut-homing cells (con 0.28±0.27, PDS 0.34±0.47, EPS±PDS 0.89±1.26, *p*=0.028 vs con, *p*=0.030 vs PDS), and increased proportions of CD8^+^ 4^+^β7^+^CCR9 cells compared to controls (0.36±0.27 vs 0.68±0.65, *p*=0.023) (**Supplementary Figure 3**). We saw no difference in gut-homing populations of the effector or memory cell populations, except for decreased CD8^+^ CM gut-homing in PDS compared to controls and EPS±PDS (con 4.11±3.19, PDS 2.29±2.68, EPS±PDS 4.60±3.91, *p*=0.028 vs con, *p*=0.011 vs EPS±PDS) (**Supplementary Figure 3**).

We used the same strategy to assess proportions of naïve, effector, CM and EM subsets in the periphery based on the expression of CD45RA, CD45RO and CCR7 in CD4^+^ (**Figure 3A**) and CD8^+^ populations (**Figure 3B**). There were no obvious shifts in the balance of PBMC effector and memory cells in FD compared to controls (**Figure 3C**) and the CD4^+^ population was unchanged (**Figure 3D**). The CD8^+^ effector population was significantly decreased in FD compared to controls (6.08±6.24 vs 2.85±2.55, *p*=0.027), attributable to PDS (2.35±63.46, *p*=0.035 vs con) (**Figure 3E**). The CD4^+^ (**Figure 3F**) and CD8^+^ (**Figure 3G**) CM populations were unaltered between controls and FD, although PDS had reduced CD8^+^ CM cells compared to EPS±PDS (1.34±1.63 vs2.71±2.22, *p*=0.007). EPS±PDS also had increased peripheral CD8^+^ naïve cells compared to controls (5.06±4.24 vs 9.23±8.56, *p*=0.041) (**Supplementary Figure 2**). These data demonstrate that while decreased CD8^+^ effector populations are observed in both FD LPMCs and PBMCs, the increase in CD4^+^ effector populations is localised to the duodenum.

**Figure 3:**
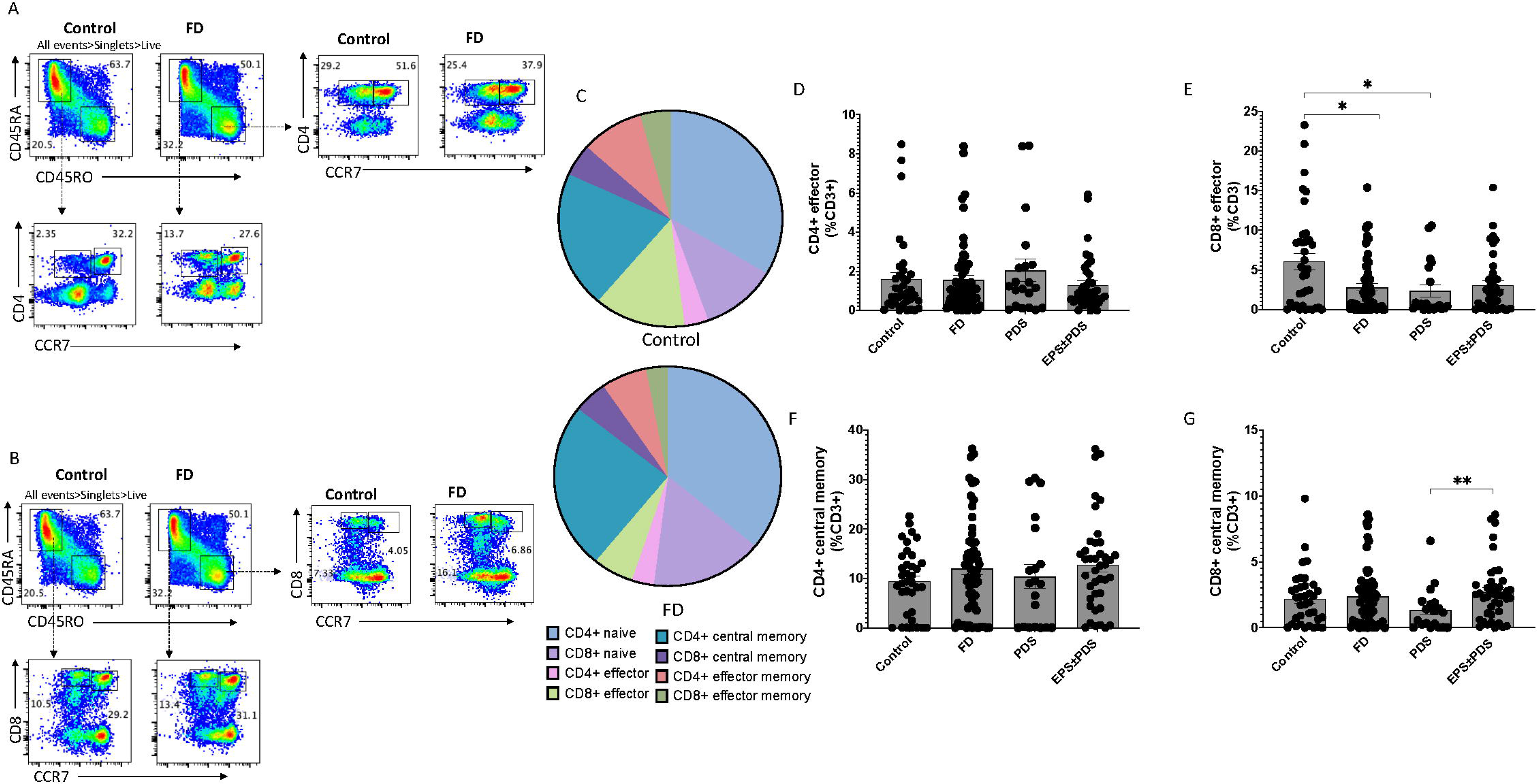
The peripheral effector and memory T cell balance in FD compared to controls. Peripheral blood mononuclear cells were isolated using density gradient centrifugation and phenotyped using surface marker staining and flow cytometry. Peripheral naïve (CD45RA^+^ CCR7^+^), effector (CD45RA^+^ CCR7^-^), central memory (CD45RO^+^ CCR7^+^) and effector memory (CD45RO^+^ CCR7^-^) T cells were identified in the (A) CD4^+^ and (B) CD8^+^ populations. (C) The balance of each effector and memory population was then represented within the total control and FD cohorts before the (D) CD4^+^ and (E) CD8^+^ effector populations were compared between controls and FD, as well as within the FD subtypes, PDS and EPS±PDS. The (F) CD4^+^ central memory and (G) CD8^+^ central memory populations were also investigated within these groups. n=37 controls, n=61 FD. Data presented as mean±SEM. Statistical analysis for control vs FD, (F) parametric t test, (D,E,G) non- parametric t test. For control vs PDS vs EPS±PDS, (F) parametric one-way ANOVA (D,E,G) non-parametric one-way ANOVA. \**p*<0.05, \*\**p*<0.01.

### Duodenal effector Th2 and Th17 populations are increased in FD patients

Th subsets have not been previously characterised in FD but are central in orchestrating specific responses of the adaptive immune system following antigen presentation. Given we have reported increased populations of duodenal CD4^+^ effector cells in FD, we used flow cytometry to characterise circulating sub-populations of Th cells. CD4^+^ effector cells were gated based on expression of CCR6, CCR4 and CXCR3 to examine the duodenal Th subsets (**Figure 4A**) and periphery (**Figure 4B**). Shifts in the effector balance were captured in pie charts in LPMC (**Figure 4C**) and PBMC populations (**Figure 4D**). There was no change in Th1 (CCR6^-^CXCR3^+^) effector (**Figure 4E**), however FD patients had increased effector Th2 (CCR6^-^CCR4^+^) cells (13.03±16.11 vs 19.84±15.51, *p*=0.038), associated with the PDS subgroup (21.22±14.58, *p*=0.04 vs con) (**Figure 4F**). In addition, effector Th17 (CCR6^+^CCR4^+^) cells were increased in FD (31.74±24.73 vs 45.57±23.75, *p*=0.03), unique to EPS±PDS (49.83±25.16, *p*=0.009 vs con) (**Figure 4G**). Conversely in the PBMCS, there was a decrease in the proportion of Th1 effector cells in FD (24.13±24.16 vs 12.70±12.74, *p*=0.049) but the Th2, Th17 and Th17.1 populations were unchanged. Interestingly, the Th1 and Th17.1 (CCR6^+^CXCR3^+^) effector populations were decreased in PDS compared to controls (**Supplementary Figure 4**).

**Figure 4:**
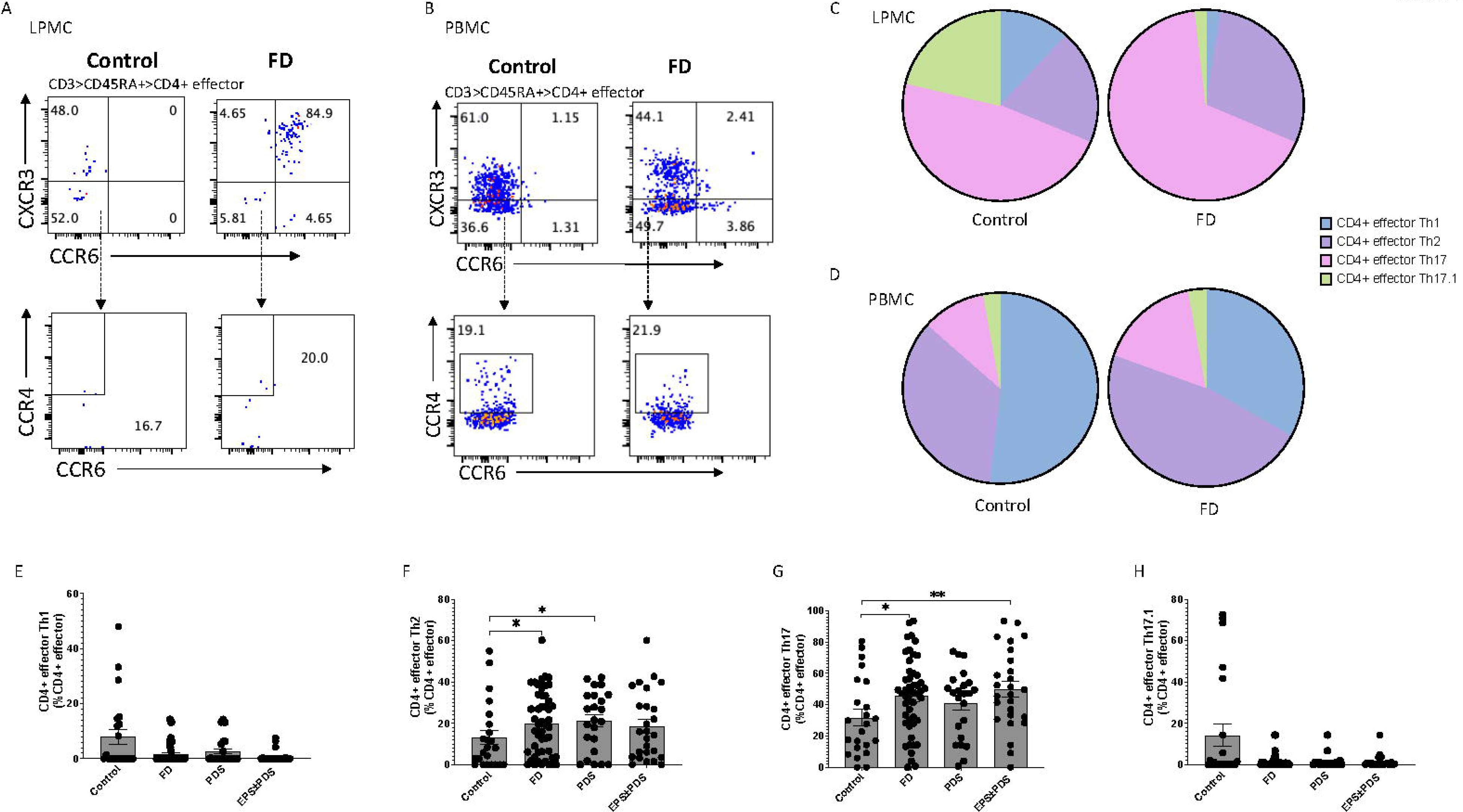
Effector T helper cells in FD patients compared to controls. Lamina propria lymphocytes were isolated from duodenal biopsies and peripheral blood mononuclear cells were isolated from whole blood. Cells were phenotyped using flow cytometry. Within the CD4^+^ effector T cell (CD4^+^ CD45RA^+^ CCR7^+^) pool, T helper cell subsets were identified based on expression of CCR6, CCR4 and CXCR3 in the isolated (A) LPMCs and (B) PBMCs. The proportions of each subset were considered in FD patients compared to controls in the (C) duodenum and (D) periphery. Within the duodenal effector populations, (E) Th1, (F) Th2, (G) Th17 and (H) Th17.1 cells were investigated in FD patients compared to controls, as well as within the FD subtypes. n=23 controls, n=49 FD for LPMCS, n=37 controls, n=61 FD for PBMCs. Data presented as mean±SEM. Statistical analysis for control vs FD, (G) parametric t test, (E,F,H) non-parametric t test. For control vs PDS vs EPS±PDS, (G) parametric and (E,F,H) non-parametric one-way ANOVA.\**p*<0.05, \*\**p*<0.01.

### Duodenal Th2 and Th17 memory T cells are also increased in FD

Given we saw a Th2/Th17 effector signature, we next investigated Th subsets in both the CM (CD45RO^+^CCR7^+^) and EM CD4^+^ populations, using the flow cytometry gating strategy outlined in the previous section on LPMCs (**Figure 5A**) and PBMCs (**Figure 5B**). Similarly to the effector populations, shifts in the proportion of each Th subset were evident in FD patients in both LPMCs (**Figure 5C**) and PBMCs (**Figure 5D**). EPS±PDS patients had decreased Th1 CM populations compared to controls (17.14±19.47 vs 9.93±10.79, *p*=0.032) (**Figure 5E**), while FD collectively had increased Th2 CM cells (23.75±18.97 vs 37.52±17.51, *p*=0.007), associated with both subtypes (PDS 35.86±19.37, *p*=0.042; EPS±PDS 38.98±16.00, *p*=0.009) (**Figure 5F**). There was no change in Th17 or Th17.1 CM populations in FD (data not shown). As the Th1/Th2 balance appeared to have shifted to a Th2 dominant signature in FD LPMCs, we examined these peripheral CM populations. In contrast to the duodenum, FD had increased proportions of CM Th1 cells (9.63±6.44 vs 15.46±11.14, *p*=0.017), attributable to PDS (17.84±13.11, *p*=0.023 vs con) (**Figure 5G**), while the peripheral CM Th2 population was unchanged (**Figure 5H**). Both the PBMC CM Th17 (31.90±19.01 vs 20.60±18.85, *p*=0.017) and Th17.1 (19.75±15.10 vs 11.59±14.76, *p*=0.026) populations were reduced in FD (**Supplementary Figure 4**).

**Figure 5:**
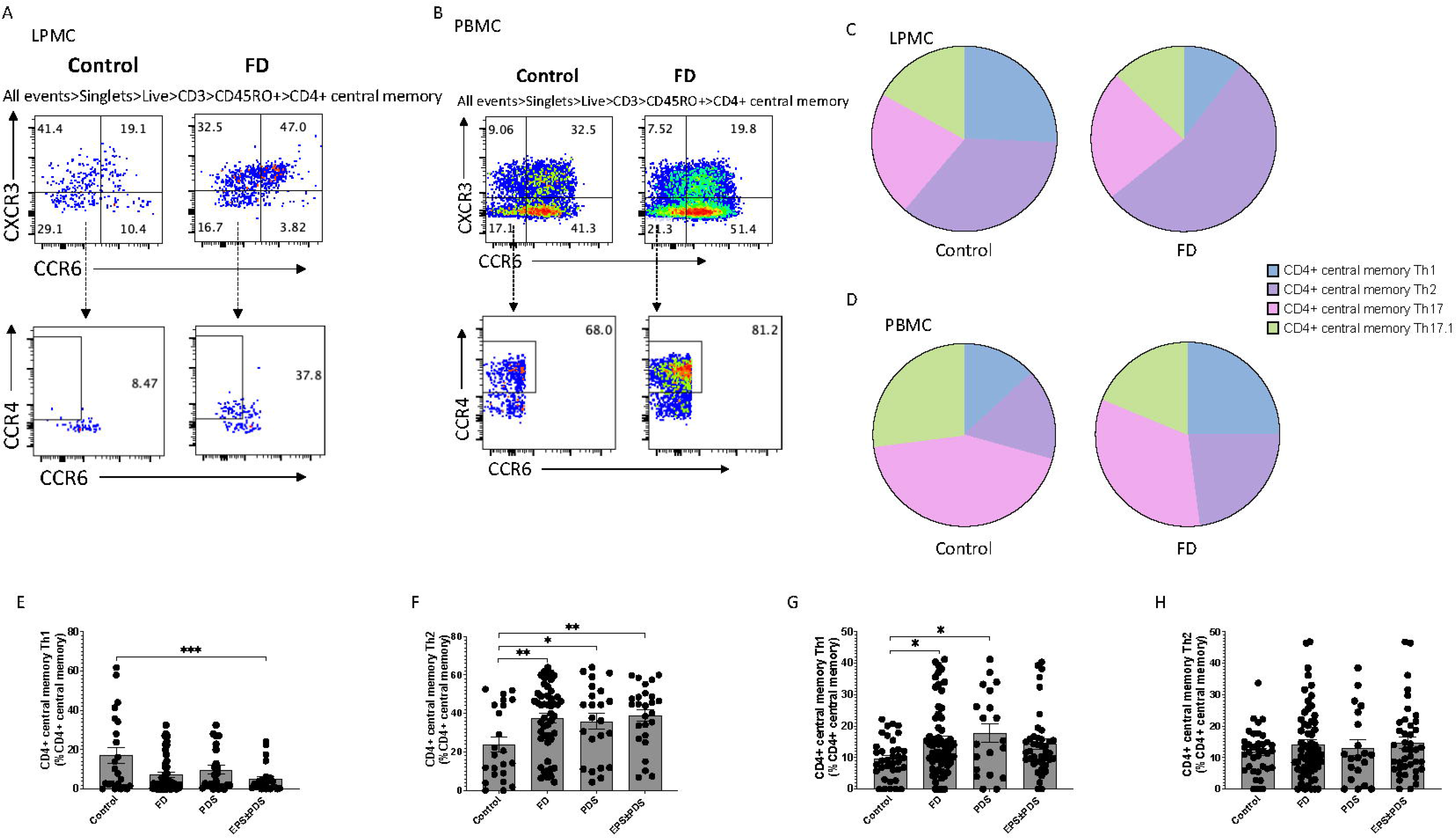
Central memory T helper cells in FD patients compared to controls. Lamina propria lymphocytes were isolated from duodenal biopsies and peripheral blood mononuclear cells were isolated from whole blood. Cells were phenotyped using flow cytometry. Within the CD4^+^ central memory T cell (CD4^+^ CD45RO^+^ CCR7^+^) pool, T helper cell subsets were identified based on expression of CCR6, CCR4 and CXCR3 in the isolated (A) LPMCs and (B) PBMCs. The proportions of each subset were considered in FD patients compared to controls in the (C) duodenum and (D) periphery. (E) Th1 and (F) Th2 central memory T cells were compared within the LPMC central memory pool and within the PBMC central memory pools ((G) Th1 and (H) Th2). n=23 controls, n=49 FD for LPMCS, n=37 controls, n=61 FD for PBMCs. Data presented as mean±SEM. Statistical analysis for control vs FD, (E,F,G,H) non-parametric t test. For control vs PDS vs EPS±PDS, (E,F,G,H) non- parametric one-way ANOVA.\**p*<0.05, \*\**p*<0.01.

We next investigated the EM (CD45RO^+^CCR7^-^) Th subpopulations in LPMCS (**Figure 6A**) and PBMCs (**Figure 6B**) and compared the proportional contribution of each subset (**Figure 6C, Figure 6D**). The Th1 EM population was unchanged (**Figure 6E**) but in line with the CM population, patients had greater proportions of Th2 EM cells (9.80±10.50 vs 20.53±14.15, *p*=0.001), common to both subtypes (PDS 22.73±16.52, *p*=0.002; EPS±PDS 18.68±11.81, *p*=0.009) (**Figure 6F**). FD also had increased proportions of EM Th17 cells (11.95±8.42 vs 18.44±15.63, *p*=0.027), specifically linked to EPS±PDS (14.39±13.15, *p*=0.018) (**Figure 6G**) while EM Th17.1 population was unchanged (**Figure 6H**). EM Th subsets were unchanged in the periphery of FD patients (**Supplementary Figure 4**).

**Figure 6:**
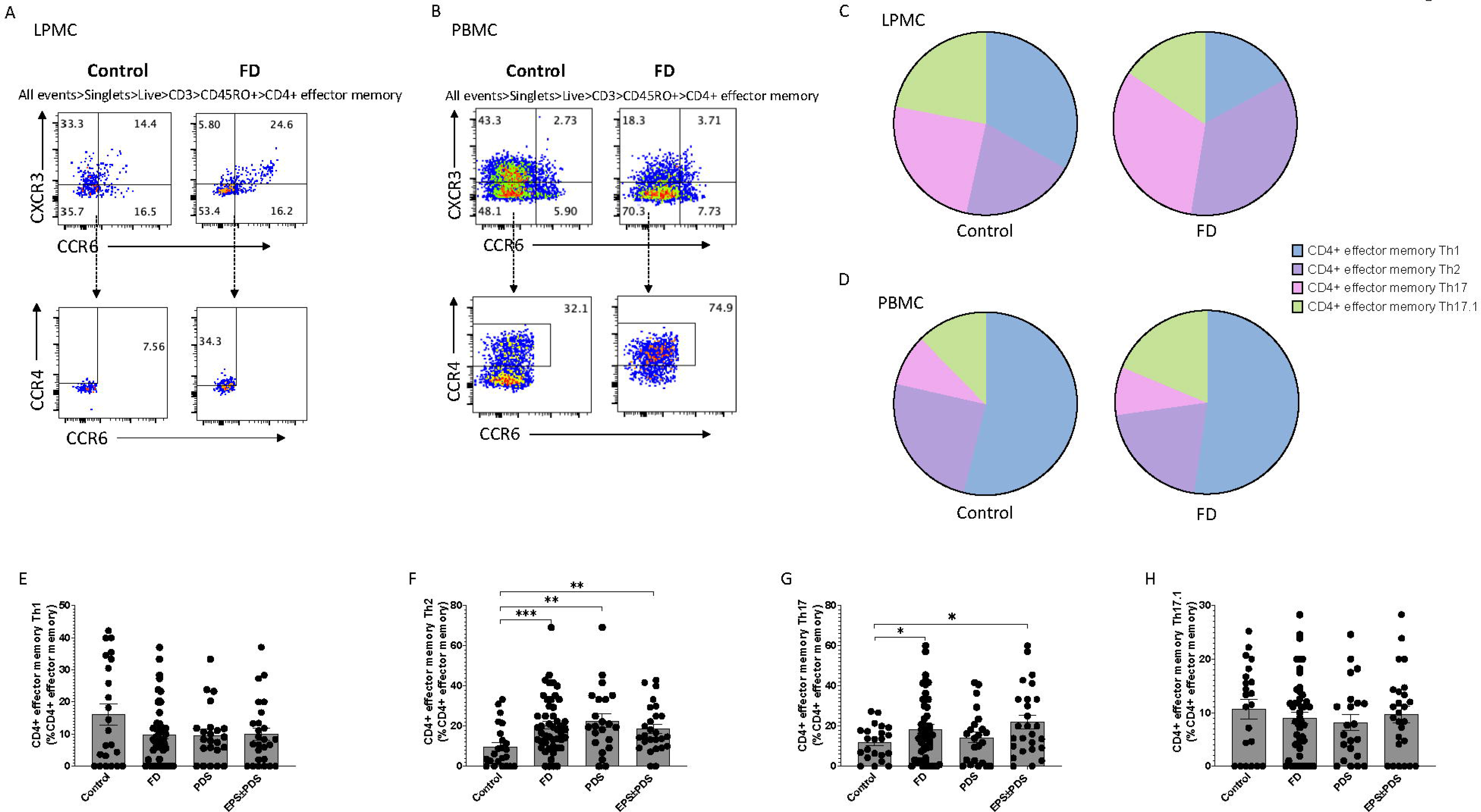
Effector memory T helper cells in FD patients compared to controls. Lamina propria lymphocytes were isolated from duodenal biopsies and peripheral blood mononuclear cells were isolated from whole blood. Cells were phenotyped using flow cytometry. Within the CD4^+^ effector memory T cell (CD4^+^ CD45RO^+^ CCR7^-^) pool, T helper cell subsets were identified based on expression of CCR6, CCR4 and CXCR3 in the isolated (A) LPMCs and (B) PBMCs. The proportions of each subset were considered in FD patients compared to controls in the (C) duodenum and (D) periphery. Within the LPMC effector memory pool, (E) Th1, (F) Th2, (G) Th17 and (H) Th17.1 populations were investigated in FD compared to controls and among the FD subtypes. n=23 controls, n=49 FD for LPMCS, n=37 controls, n=61 FD for PBMCs. Data presented as mean±SEM. Statistical analysis for control vs FD, (G,H) parametric t test, (E,F) non-parametric t test. For control vs PDS vs EPS±PDS, (G,H) parametric and (E,F) non-parametric one-way ANOVA.\**p*<0.05, \*\**p*<0.01.

Collectively, these data demonstrate the effector and memory Th1/Th2 balances are altered in FD compared to controls, with patients demonstrating a predominant Th2 and Th17 phenotype. Further, these data demonstrate that unstimulated PBMC populations do not reflect the duodenal balance in FD.

### Cohort characteristics have minimal influence on T cell profiles

We also compared T cell profiles in the duodenum (**Supplementary Table 3**) and periphery (**Supplementary Table 4**) of FD with (FD+IBS) and without concomitant IBS, given the presence of multiple DGBIs is associated with greater symptom severity.[16] There was no difference in the proportions of CD4^+^ (**Figure 7A**), CD8^+^ (**Figure 7B**) gut-homing or CD4^+^ effector cells (**Figure 7C**) in FD compared to FD+IBS. With regards to the CD4^+^ Th subsets, there were no difference in the effector Th1 (**Figure 7D**) or Th2 (**Figure 7E**), CM Th1 (**Figure 7F**) or Th2 (**Figure 7G**) subsets. There was no change in EM Th1 (**Figure 7H**) but the EM Th2 population was increased in FD+IBS compared to FD without IBS (17.85±11.50 vs 26.45±17.76, *p*=0.034) (**Figure 7I**). These data suggest concomitant IBS and FD is associated with a greater proportion of EM Th2 cells compared to FD only.

**Figure 7:**
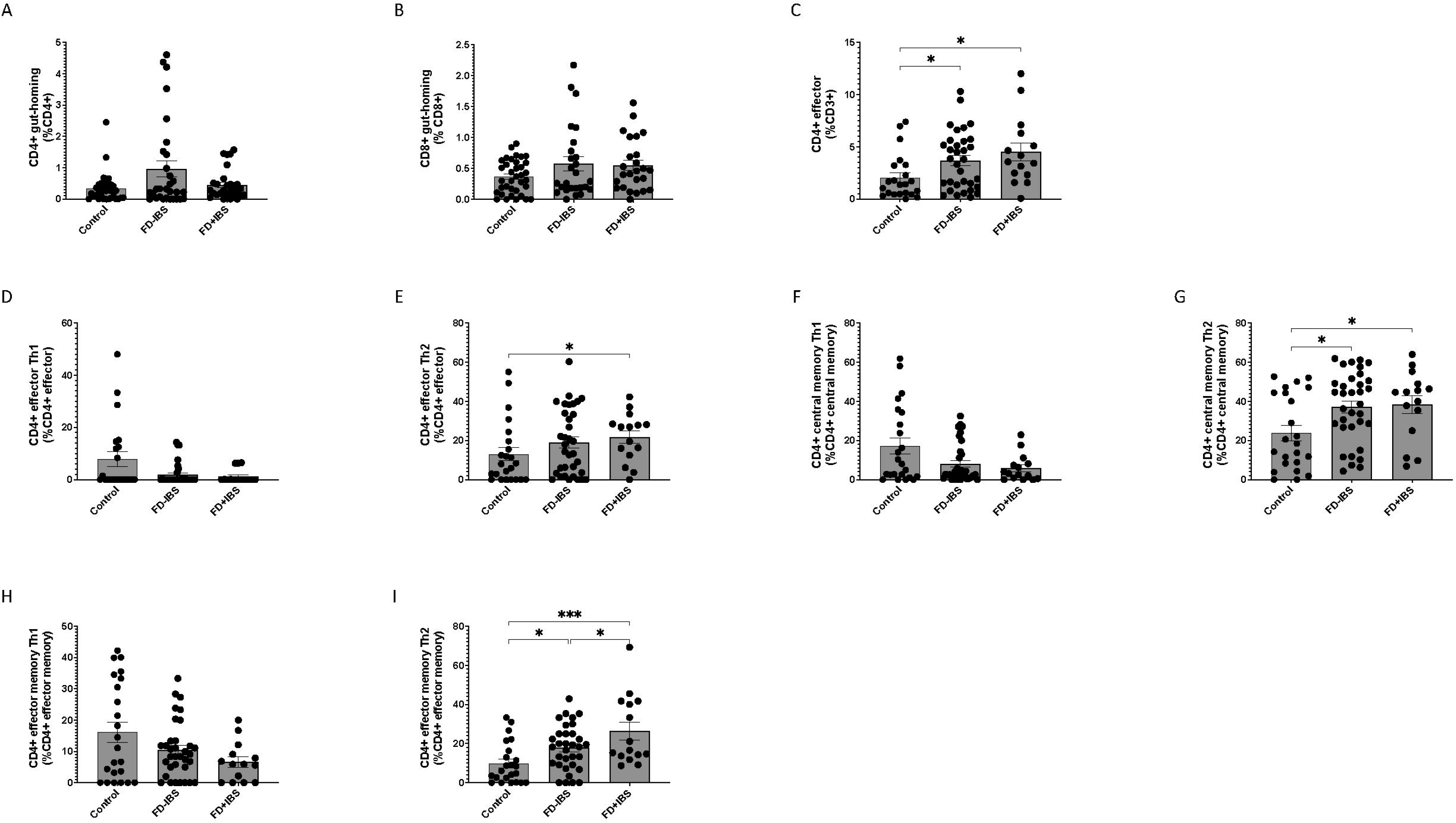
T cell profiles in FD patients with and without concomitant IBS. The duodenal and peripheral T cell profiles generated using flow cytometry were analysed in the context of FD patients compared to FD patients with concomitant IBS (FD+IBS). The proportions of (A) peripheral CD4^+^ gut-homing, (B) peripheral CD8^+^ gut-homing and (C) duodenal CD4^+^ effector T cells were compared between groups. The duodenal Th subsets were also examined in the effector ((D) Th1 and (E) Th2), central memory ((F) Th1, (G) Th2) and effector memory ((H) Th1, (I) Th2) populations in FD±IBS. n=23 controls, n= FD, n= FD+IBS. Data presented as mean±SEM. Statistical analysis (I) parametric and (A,B,C,D,E,F,G,H) non-parametric one-way ANOVA. \**p*<0.05, \*\*\**p*<0.001.

Given there were differences in our cohort regarding factors including PPI usage **(Supplementary Tables 5 and 6**) and sex **(Supplementary Tables 7 and 8**), we investigated the influence of these factors on duodenal and peripheral profiles. Interestingly, we observed that FD patients not on PPIs had increased duodenal EM and CM Th2 proportions compared to controls, and that the CM Th17 population was increased compared to FD+PPI. These findings support the notion that PPIs have immunomodulatory capacity in the duodenum.

### Cytokine activity in FD

While several studies reported alterations in cytokine populations in FD,[15] few studies have looked at cytokines within Rome subtypes, or compared circulating cytokines with cytokines released from duodenal LPMCs. Plasma and duodenal supernatants were assayed for cytokine levels where available. Plasma IL-10 was decreased in FD (40.68±7.850pg/mL vs 32.13±7.905pg/mL, *p*=0.003), and in subtypes (PDS 32.68±5.177pg/mL, *p*=0.031 vs controls, EPS±PDS 31.80±9.279pg/mL, *p*=0.029 vs controls) (**Figure 8A**). There was no change in duodenal supernatant IL-10 (**Figure 8B**). While plasma TNF was decreased in FD (56.85±12.16pg/mL vs 48.22±9.646pg/mL, *p*=0.032), specifically PDS (PDS 44.85±11.03pg/mL, *p*=0.007 vs controls, EPS±PDS 50.30±8.234pg/mL, *p*=0.146 vs controls) (**Figure 8C**), there was no difference in supernatant TNF levels (**Figure 8D**). Plasma IFN-γ was decreased in FD (64.04±10.08pg/mL vs 55.10±8.285pg/mL *p*=0.01) and PDS (PDS 51.60±7.025pg/mL, *p*=0.0007 vs controls, EPS±PDS 57.25±8.388pg/mL, *p*=0.056 vs controls) (**Figure 8E**). Assessment of duodenal IFN-γ (**Figure 8F**), IL-17a levels in plasma (**Figure 8G**) and supernatants (**Figure 8H**) revealed no change between controls and FD.

**Figure 8:**
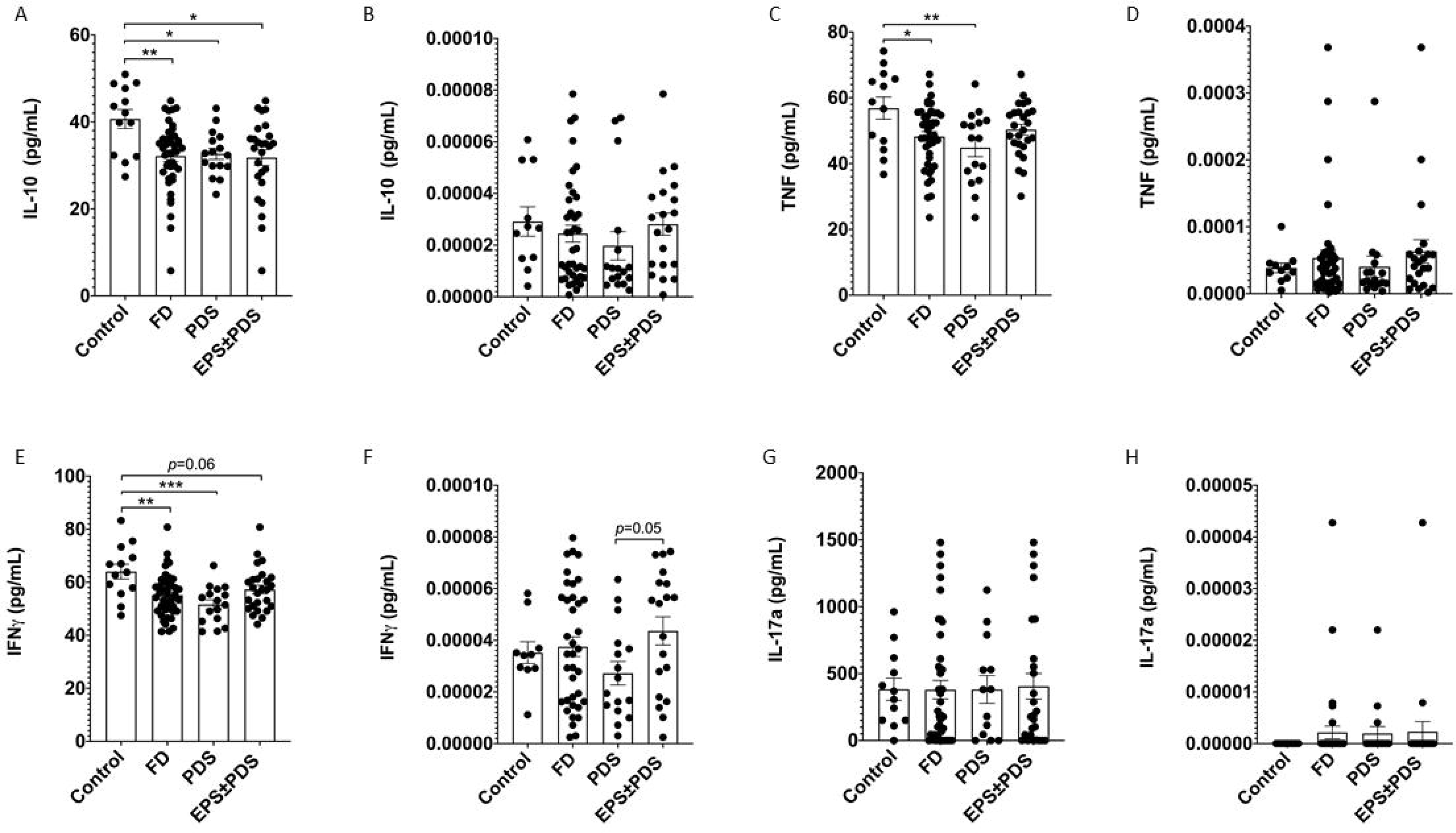
Plasma and LPMC supernatant cytokine levels (pg/mL) in controls and FD patients. Plasma collected during the PBMC isolation process, and media supernatants collected from LPMC cells incubated overnight were assayed for cytokine expression using a CBA kit for Th1/Th2/Th17 cytokines. IL-10 levels were measured in the (A) plasma and (B) LPMC supernatant of controls, FD, PDS and EPS±PDS patients. TNF was also investigated in (C) plasma and (D) supernatants collected from controls, FD, PDS and EPS±PDS subjects. The level of IFN-γ was assayed in (E) plasma and (F) supernatants from controls, FD, PDS and EPS±PDS patients. IL-17a concentrations were assayed in (G) plasma and (H) supernatants in controls, FD, PDS and EPS±PDS subjects. n=12-13 controls, n=39 patients. Data presented as mean±SEM. Statistical analysis for control vs FD, (C,E) parametric and (A,B,D,F,G,H) non-parametric t test. For control vs PDS vs EPS±PDS, (C,E,F,G) parametric and (A,B,D,H) non-parametric one-way ANOVA.*p<0.05,**p<0.01,***p<0.001.

## DISCUSSION

Imnune activation may play a role in symptom manifestation of patients with FD and IBS. However to date, while some studies have used selected markers to capture mucosal or systemic activation of the immune system, no comprehensive assessment of the T cell phenotype in FD has been performed.[15] As such, this study aimed to phenotype lymphocyte populations in the mucosa and periphery of FD patients to identify those involved in the characteristic mucosal dysregulation. Our findings have revealed a duodenal Th2 and Th17 signature in FD compared to outpatient controls, suggesting symptom onset may be associated with stimulation of antigen-experienced T cells. Consistent with previous work, and supporting the notion of FD as a disorder of homeostatic imbalance,[9, 12] we did not find alterations in the proportions of CD3^+^, CD4^+^ or CD8^+^ lymphocytes. Instead, our findings suggest FD exhibits a Th2/Th17 response localised to the duodenum. Interestingly, the duodenal phenotype was not observed in the PBMC profiles of patients, further supporting recent work demonstrating that localised responses to food antigens activate the mucosal immune response in both FGID patients and animal models of visceral hyspersensitivity.[20, 21] Such discrepancies between the systemic and mucosal response suggest caution is warrented when interpreting immune profiles from unstimulated PBMCs in these conditions. Given that sourcing PBMCs is less invasive than obtaining LPMCs, one important question to be answered is whether the memory phenotype from the localised responses can be provoked in PBMCs using luminal antigen stimulation, thus overcoming the need for biopsy-derived lymphocytes to verify the FD immunophenotype.

Given Th2 immune responses are associated with eosinophil recruitment and activation[22], our findings would suggest provocation of the immune response by a specific antigen may be associated with duodenal eosinophilia. We also identified increased proportions of Th17 cells in the duodenum, a Th subset traditionally associated with autoimmune conditions, including Crohn’s disease[23], coeliac disease[24] and rheumatoid arthritis[25]; or the adaptive immune response to extracellular pathogens.[26] The concept of multiple Th subsets activated in the mucosa has a precedence in asthma, where inflammatory signatures have been implicated in simultaneous activation of Th17 and Th2 responses.[27] An existing theory is that these overlapping responses are the result of multiple environmental antigen exposures, such as a concurrent infection and exposure to allergen; or that these responses represent generation of autoimmune reactions due to chronic cycles of inflammation and repair.[28] This scenario is plausible in FD, given that FD can develop after acute gastroenteritis, and is also associated with both atopic and autoimmune conditions.[29, 30, 31] Another hypothesis from asthma studies suggests Th17 cells are activated where Th2 responses are ineffective in response to an infection or antigen.[27] Th17 cells then regulate Th2 cells to restore homeostasis,[32] but failure to resolve the response may drive cyclic symptoms. As such, we would propose a multi-antigen model for chronic immune activation in FD. In this hypothetical scenario, disruption to duodenal homeostasis (such as infection) would drive physiological disruption to the GI tract and allow for greater contact between mucosal immune cells and luminal antigens. Once the stimulus is removed and the immune response lapses, Th2 and Th17 memory cells remain in the duodenum for future exposures.

Our examination of cytokine levels in the plasma and unstimulated LPMC supernatants revealed that peripheral concentrations of IL-10, TNF and IFN-γ were significantly decreased compared to controls, and IL-17a levels were unchanged. We have previously identified discrepancies in reported levels of cytokines in FGID studies, due to methodological differences.[15] However, given the brief half-lives of cytokines and physiological influences on their concentrations,[33, 34] it is plausible that sampling blood and biopsies at time of endoscopy, and thus in the absence of antigenic stimulation, does not capture the active response, given the cyclic nature of symptom onset. As such, we interpret these cytokine levels with extreme caution and suggest that the lack of a known antigen to induce a specific change in cytokine levels does not exclude these populations from involvement in FD. Unfortunately, this will remain a significant limitation to similar studies in this field until specific antigens can be identified.

Our study is limited by the small number of controls and by their outpatient status. The ideal phenotyping study would compare to healthy controls, however ethical considerations in obtaining matched biopsies complicate this. However, we believe careful characterisation of both our controls and FD cohort for organic diseases and immune confounders (including BMI and age) has mitigated this limitation as much as possible. While some of our cohort had overlapping IBS, these patients were recruited to the study only where their FD symptoms were the primary complaint. Further, the duodenal Th2 and Th17 signatures we observed in FD were not specific to the overlap cohort. While we have been able to identify significant differences in cell populations between FD and controls, we believe our cohort is underpowered to effectively examine these cell populations under the Rome III criteria subtypes. We had small numbers of ‘pure’ EPS subjects, as most reported both EPS and PDS symptoms, and it is recognised that post-prandial epigastric pain largely contributes to this overlap.[2, 35] Of note, this study used Rome III for consistency, as recruitment commenced prior to Rome IV but the updated criteria are similar.[36]

The findings of this study demonstrate that FD patients have increased proportions of specific effector T cells in the duodenum, confirming immune activation is a feature of FD. Further, for the first time, we have shown patients have increased duodenal Th2 and Th17 cells, confined to the duodenum. Importantly neither Rome criteria subgrouping nor IBS overlap account for this immunophenotype in our cohort. Our findings suggest dual lymphocyte response pathways are involved in FD symptom generation, giving new insights into the aetiology of this condition.

## AUTHOR CONTRIBUTIONS

S.K., N.J.T., M.M.W. and G.H participated in the design of the concept, hypothesis, and aims of the study. G.L.B and S.K participated in initial drafting of the manuscript. G.L.B performed sample processing, immunophenotyping experiments, cytokine assays and analysis. J.B., A.M., K.M., and T.F. assisted with collection and processing of patient samples. G.L.B, R.C. and M.M.W. performed histological analysis.

P.M.N. advised on data analysis. M.D.E.P, S.B., M.Z.I., L.T.G., R.F., M.V., A.S., and N.J.T. assisted with recruitment and review of cohort. P.S.F and J.H. assisted with resources, experimental design and manuscript editing. C.N., N.P., N.J.T, M.M.W assisted with concept, experimental design and manuscript editing. All the authors read and approved the final manuscript.

## Data Availability

All data produced in the present work are contained in the manuscript

## ACKNOWLEDGEMENTS

The authors would like to acknowledge the support of Nicole Cole (Analytical Biomolecular Research Facility, University of Newcastle, Australia) as well as Dr. Andrew Lim, Dr. Carole Ford, Dr. Nikki Alling, Dr. Frank Kao and Dr. Ali Kamene (BD Biosciences, Australia and New Zealand) for their help and assistance with flow cytometry for this work. We thank the Hunter Medical Research Institute Core Histology Facility for processing, sectioning and staining histological sections, and Megan Clarke (Hunter Cancer Biobank/NSW Regional Biopsecimen & Research Services) for immunohistochemical staining. We also would like to thank Dr. Sherine Hermangild Kottoor from the Powell Lab (Imperial College London, London, United Kingdom) for insightful conversations around how to approach the phenotypic analysis.

## SUPPLEMENTARY FIGURE LEGENDS

**Supplementary Figure 1: Duodenal and peripheral lymphocytes in functional dyspepsia** Lamina propria lymphocytes were isolated from duodenal biopsies and peripheral blood mononuclear cells were isolated from whole blood. Cells were phenotyped using flow cytometry. In the LPMCS, (A) CD3^+^, (B) CD4^+^ and (C) CD8^+^ populations were investigated, along with the PBMC (D) CD3^+^, (E) CD4^+^ and (F) CD8^+^ populations in FD patients compared to controls, as well as within the FD subtypes. n=23 controls, n=49 FD for LPMCS, n=37 controls, n=61 FD for PBMCs. Data presented as mean±SEM. Statistical analysis for control vs FD, (A,B,C,D,E) parametric t test, (F) non-parametric t test. For control vs PDS vs EPS±PDS, (A,B,D,E) parametric one-way ANOVA, (C,F) non-parametric one-way ANOVA. \**p*<0.05.

**Supplementary Figure 2: Duodenal and peripheral naïve and effector memory T cell populations in FD patients compared to controls.** Lamina propria mononuclear cells were isolated from duodenal biopsies and phenotyped using surface marker staining and flow cytometry. (A) CD4^+^ and (B) CD8^+^ naïve (CD45RA^+^ CCR7^+^), along with (C) CD4^+^ and (D) CD8^+^ effector memory (CD45RO^+^ CCR7^-^) T cell populations were compared between controls and FD patients, as well as within the PDS and EPS±PDS subtypes of FD. Peripheral blood mononuclear cells were isolated using density gradient centrifugation and phenotyped using surface marker staining and flow cytometry as described above. (E) CD4^+^ and (F) CD8^+^ naïve (CD45RA^+^ CCR7^+^), along with (G) CD4^+^ and (H) CD8^+^ effector memory (CD45RO^+^ CCR7^-^) T cell populations were compared between controls and FD patients, as well as within the PDS and EPS±PDS subtypes of FD. n=23 controls, n=49 FD for LPMCS, n=37 controls, n=61 FD for PBMCs. Data presented as mean±SEM. Statistical analysis for control vs FD, (A,B,C,D,E,F,G,H) non-parametric t test. For control vs PDS vs EPS±PDS, (A,B,C,D,E,F,G,H) non-parametric one-way ANOVA.

**Supplementary Figure 3: Gut-homing T cells in the periphery of FD patients compared to controls** Peripheral blood mononuclear cells were isolated from whole blood and the proportions of (A) CD4^+^ 4^+^ 7^+^ CCR9^+^ cells were enumerated in (B) controls, FD, PDS and EPS±PDS groups. (C) Populations of CD8^+^ 4^+^ 7^+^ CCR9^+^ cells were also investigated in (D) control, FD, PDS and EPS±PDS groups. Within the CD3^+^ T cell pool, populations of gut-homing T cells were investigated in (E) CD4^+^ and (F) CD8^+^ effector (CD45RA^+^ CCR7^-^), (G) CD4^+^ and (H) CD8^+^ central memory (CD45RO^+^ CCR7^+^) and the (I) CD4^+^ and (J) CD8^+^ effector memory (CD45RO^+^ CCR7^-^) populations. n=37 controls, n=61 FD. Data presented as mean±SEM. Statistical analysis for control vs FD, (E,F,G,H,I,J) non-parametric *t* test. For control vs PDS vs EPS±PDS, (E,F,G,H,I,J) non-parametric one-way ANOVA. Non- parametric *t* tests with one-tailed *p* value (B, D).\**p*<0.05.

**Supplementary Figure 4: T helper subsets in FD patients compared to controls.** Peripheral blood mononuclear cells were isolated from whole blood. Cells were phenotyped using flow cytometry. Within the CD4^+^ effector T cell (CD45RA^+^ CCR7^+^), central memory (CD45RO^+^ CCR7^+^) and effector memory CD45RO^+^ CCR7^-^) pools, T helper cell subsets were identified based on expression of CCR6, CCR4 and CXCR3. Within the peripheral effector populations, (A) Th1, (B) Th2, (C) Th17 and (D) Th17.1 cells were investigated in FD patients compared to controls, as well as within the FD subtypes. (E) Th17 and (F) Th17.1 central memory and (G) Th1, (H) Th2, (I) Th17 and (Th17.1) effector memory T cell populations were also investigated in this cohort. n=37 controls, n=61 FD for PBMCs. Data presented as mean±SEM. Statistical analysis for control vs FD, (H) parametric t test, (A,B,C,D,E,F,G,I,J) non-parametric t test. For control vs PDS vs EPS±PDS, (D,H) parametric and (A,B,C,E,F,G,I,J) non-parametric one-way ANOVA.\**p*<0.05.

## REFERENCES

1. Drossman DA. Functional Gastrointestinal Disorders: History, Pathophysiology, Clinical Features and Rome IV. Gastroenterology 2016;150:1262–79.e2.

2. Carbone F, Vanuytsel T, Tack J. Analysis of Postprandial Symptom Patterns in Subgroups of Patients With Rome III or Rome IV Functional Dyspepsia. Clinical Gastroenterology and Hepatology 2020;18:838-+.

3. Walker MM, Talley NJ, Prabhakar M, Pennaneac’h CJ, Aro P, Ronkainen J, et al. Duodenal mastocytosis, eosinophilia and intraepithelial lymphocytosis as possible disease markers in the irritable bowel syndrome and functional dyspepsia. Aliment Pharmacol Ther 2009;29:765–73.

4. Talley NJ, Walker MM, Aro P, Ronkainen J, Storskrubb T, Hindley LA, et al. Non- ulcer dyspepsia and duodenal eosinophilia: an adult endoscopic population-based case- control study. Clin Gastroenterol Hepatol 2007;5:1175–83.

5. Walker MM, Salehian SS, Murray CE, Rajendran A, Hoare JM, Negus R, et al. Implications of eosinophilia in the normal duodenal biopsy - an association with allergy and functional dyspepsia. Aliment Pharmacol Ther 2010;31:1229–36.

6. Vanheel H, Vicario M, Vanuytsel T, Van Oudenhove L, Martinez C, Keita AV, et al. Impaired duodenal mucosal integrity and low-grade inflammation in functional dyspepsia. Gut 2014;63:262–71.

7. Walker MM, Aggarwal KR, Shim LSE, Bassan M, Kalantar JS, Weltman MD, et al. Duodenal eosinophilia and early satiety in functional dyspepsia: Confirmation of a positive association in an Australian cohort. Journal of Gastroenterology and Hepatology 2014;29:474–9.

8. Wang X, Li X, Ge W, Huang J, Li G, Cong Y, et al. Quantitative evaluation of duodenal eosinophils and mast cells in adult patients with functional dyspepsia. Ann Diagn Pathol 2015;19:50–6.

9. Liebregts T, Adam B, Bredack C, Gururatsakul M, Pilkington KR, Brierley SM, et al. Small bowel homing T cells are associated with symptoms and delayed gastric emptying in functional dyspepsia. Am J Gastroenterol 2011;106:1089–98.

10. Ishigami H, Matsumura T, Kasamatsu S, Hamanaka S, Taida T, Okimoto K, et al. Endoscopy-Guided Evaluation of Duodenal Mucosal Permeability in Functional Dyspepsia. Clin Transl Gastroenterol 2017;8:e83.

11. Komori K, Ihara E, Minoda Y, Ogino H, Sasaki T, Fujiwara M, et al. The Altered Mucosal Barrier Function in the Duodenum Plays a Role in the Pathogenesis of Functional Dyspepsia. Dig Dis Sci 2019;64:3228–39.

12. Kindt S, Van Oudenhove L, Broekaert D, Kasran A, Ceuppens JL, Bossuyt X, et al. Immune dysfunction in patients with functional gastrointestinal disorders. Neurogastroenterology and Motility 2009;21:389–98.

13. Farrar JD, Asnagli H, Murphy KM. T helper subset development: roles of instruction, selection, and transcription. Journal of Clinical Investigation 2002;109:431–5.

14. Keely S, Foster PS. Stop Press: Eosinophils Drafted to Join the Th17 Team. Immunity 2015;43:7–9.

15. Burns G, Carroll G, Mathe A, Horvat J, Foster P, Walker MM, et al. Evidence for Local and Systemic Immune Activation in Functional Dyspepsia and the Irritable Bowel Syndrome: A Systematic Review. Am J Gastroenterol 2019;114:429–36.

16. Von Wulffen M, Talley NJ, Hammer J, McMaster J, Rich G, Shah A, et al. Overlap of Irritable Bowel Syndrome and Functional Dyspepsia in the Clinical Setting: Prevalence and Risk Factors. Digestive Diseases and Sciences 2019;64:480–6.

17. Corsetti M, Caenepeel P, Fischler B, Janssens J, Tack J. Impact of coexisting irritable bowel syndrome on symptoms and pathophysiological mechanisms in functional dyspepsia. Am J Gastroenterol 2004;99:1152–9.

18. Talley NJ, Haque M, Wyeth JW, Stace NH, Tytgat GN, Stanghellini V, et al. Development of a new dyspepsia impact scale: the Nepean Dyspepsia Index. Aliment Pharmacol Ther 1999;13:225–35.

19. Marks E, Naudin C, Nolan G, Goggins BJ, Burns G, Mateer SW, et al. Regulation of IL-12p40 by HIF controls Th1/Th17 responses to prevent mucosal inflammation. Mucosal Immunology 2017;10:1224–36.

20. Aguilera-Lizarraga J, Florens MV, Viola MF, Jain P, Decraecker L, Appeltans I, et al. Local immune response to food antigens drives meal-induced abdominal pain. Nature 2021.

21. Fritscher-Ravens A, Pflaum T, Mosinger M, Ruchay Z, Rocken C, Milla PJ, et al. Many Patients With Irritable Bowel Syndrome Have Atypical Food Allergies Not Associated With Immunoglobulin E. Gastroenterology 2019;157:109–18 e5.

22. Paul WE, Zhu J. How are T(H)2-type immune responses initiated and amplified? Nat Rev Immunol 2010;10:225–35.

23. Brand S. Crohn’s disease: Th1, Th17 or both? The change of a paradigm: new immunological and genetic insights implicate Th17 cells in the pathogenesis of Crohn’s disease. Gut 2009;58:1152–67.

24. Mazzarella G. Effector and suppressor T cells in celiac disease. World J Gastroenterol 2015;21:7349–56.

25. Kamali AN, Noorbakhsh SM, Hamedifar H, Jadidi-Niaragh F, Yazdani R, Bautista JM, et al. A role for Th1-like Th17 cells in the pathogenesis of inflammatory and autoimmune disorders. Mol Immunol 2019;105:107–15.

26. Uchiyama R, Yonehara S, Taniguchi S, Ishido S, Ishii KJ, Tsutsui H. Inflammasome and Fas-Mediated IL-1beta Contributes to Th17/Th1 Cell Induction in Pathogenic Bacterial Infection In Vivo. J Immunol 2017;199:1122–30.

27. Ramakrishnan RK, Al Heialy S, Hamid Q. Role of IL-17 in asthma pathogenesis and its implications for the clinic. Expert Rev Respir Med 2019;13:1057–68.

28. Erjefalt JS. Unravelling the complexity of tissue inflammation in uncontrolled and severe asthma. Curr Opin Pulm Med 2019;25:79–86.

29. Koloski N, Jones M, Walker MM, Veysey M, Zala A, Keely S, et al. Population based study: atopy and autoimmune diseases are associated with functional dyspepsia and irritable bowel syndrome, independent of psychological distress. Aliment Pharmacol Ther 2019;49:546–55.

30. Ford AC, Talley NJ, Walker MM, Jones MP. Increased prevalence of autoimmune diseases in functional gastrointestinal disorders: case-control study of 23471 primary care patients. Aliment Pharmacol Ther 2014;40:827–34.

31. Walker MM, Talley NJ, Keely S. Follow up on atopy and the gastrointestinal tract - a review of a common association 2018. Expert Rev Gastroent 2019;13:437–45.

32. Ciofani M, Madar A, Galan C, Sellars M, Mace K, Pauli F, et al. A validated regulatory network for Th17 cell specification. Cell 2012;151:289–303.

33. Zhou X, Fragala MS, McElhaney JE, Kuchel GA. Conceptual and methodological issues relevant to cytokine and inflammatory marker measurements in clinical research. Curr Opin Clin Nutr Metab Care 2010;13:541–7.

34. Friberg D, Bryant J, Shannon W, Whiteside TL. In-Vitro Cytokine Production by Normal Human Peripheral-Blood Mononuclear-Cells as a Measure of Immunocompetence or the State of Activation. Clin Diagn Lab Immun 1994;1:261–8.

35. Carbone F, Holvoet L, Tack J. Rome III functional dyspepsia subdivision in PDS and EPS: recognizing postprandial symptoms reduces overlap. Neurogastroenterol Motil 2015;27:1069–74.

36. Schmulson MJ, Drossman DA. What Is New in Rome IV. J Neurogastroenterol Motil 2017;23:151–63.

