## Supplementary information for "Distinct Adaptive Immunophenotypes in duodenal mucosa but not in peripheral blood of patients with functional dyspepsia"

Supplementary Table 1: List of antibodies used in surface marker flow cytometry staining for identification of lymphocyte subsets

| **Antibody target** | **Fluorophore** | **Clone** | **Supplier** | **Vol./100µL test** |
|---|---|---|---|---|
| CD3 | BUV805 | UCHT1 | BD Biosciences, cat no. 612896 | 2µL |
| CD4 | FITC | RPA-T4 | BD Biosciences, cat no. 555346 | 5µL |
| CD8 | BUV496 | RPA-T8 | BD Biosciences, cat no. 612942 | 2µL |
| CD45RA | BUV395 | 5H9 | BD Biosciences, cat no. 740315 | 2µL |
| CD45RO | PE-CY7 | UCHL1 | BD Biosciences, cat no. 560608 | 2µL |
| CCR7 (CD197) | BV711 | 150503 | BD Biosciences, cat no. 566602 | 5µL |
| CCR6 (CD196) | BV786 | 11A9 | BD Biosciences, cat no. 563704 | 5µL |
| CCR4 (CD194) | BV421 | 1G1 | BD Biosciences, cat no. 562579 | 5µL |
| CXCR3 (CD183) | PE | 1C6 | BD Biosciences, cat no. 557185 | 15µL |
| Integrin α4 | PE-CF594 | 9F10 | BD Biosciences, cat no. 563645 | 5µL |
| Integrin β7 | BV650 | FIB504 | BD Biosciences, cat no. 564285 | 5µL |
| CCR9 | APC | 112509 | BD Biosciences, cat no. 557975 | 2µL |
| Fixable viability dye | FVS700 | N/A | BD Biosciences, cat no. 564997 | 1:1000 |

Supplementary Table 2: Surface markers for the identification of T cell subsets

| T cell subset | Surface maker phenotype |
|---|---|
| Lymphocytes | $CD3^{+}$ |
| T helper cells | $CD3^{+}CD4^{+}$ |
| Cytotoxic T cells | $CD3^{+}CD8^{+}$ |
| Naïve T cells | $CD4^{+}CD45RA^{+}CCR7^{+}/CD8^{+}CD45RA^{+}CCR7^{+}$ |
| Effector T cells | $CD4^{+}CD45RA^{+}CCR7^{-}/CD8^{+}CD45RA^{+}CCR7^{-}$ |
| Central memory T cells | $CD4^{+}CD45RO^{+}CCR7^{+}/CD8^{+}CD45RO^{+}$ $CCR7^{+}$ |
| Effector memory T cells | $CD4^{+}CD45RO^{+}CCR7^{-}/CD8^{+}CD45RO^{+}CCR7^{-}$ |
| Small intestinal homing T cells | $CD4^{+}\alpha4^{+}\beta7^{+}CCR9^{+}/CD8^{+}\alpha4^{+}\beta7^{+}CCR9^{+}$ |
| Antigen-experienced T cells | $CD3^{+}CD4^{+}CD45RO^{+}$ |
| T helper 1 T cells | $CD3^{+}CD4^{+}CD45RO^{+}CCR6^{-}CXCR3^{+}$ |
| T helper 2 T cells | $CD3^{+}CD4^{+}CD45RO^{+}CCR6^{-}CCR4^{+}$ |
| T helper 17 T cells | $CD3^{+}CD4^{+}CD45RO^{+}CCR6^{+}CCR4^{+}$ |
| T helper 17.1 T cells | $CD3^{+}CD4^{+}CD45RO^{+}CCR6^{+}CXCR3^{+}$ |
| Effector Th1 T cells | $CD4^{+}CD45RA^{+}CCR7^{-}CCR6^{-}CXCR3^{+}$ |
| Effector Th2 T cells | $CD4^{+}CD45RA^{+}CCR7^{-}CCR6^{-}CCR4^{+}$ |
| Effector Th17 T cells | $CD4^{+}CD45RA^{+}CCR7^{-}CCR6^{+}CCR4^{+}$ |
| Effector Th17.1 T cells | $CD4^{+}CD45RA^{+}CCR7^{-}CCR6^{+}CXCR3^{+}$ |
| Effector gut-homing T cells | $CD4^{+}CD45RA^{+}CCR7^{-}\alpha4^{+}\beta7^{+}CCR9^{+}/CD8^{+}CD45RA^{+}CCR7^{-}\alpha4^{+}\beta7^{+}CCR9^{+}$ |
| Central memory Th1 T cells | $CD4^{+}CD45RO^{+}CCR7^{+}CCR6^{-}CXCR3^{+}$ |
| Central memory Th2 T cells | $CD4^{+}CD45RO^{+}CCR7^{+}CCR6^{-}CCR4^{+}$ |
| Central memory Th17 T cells | $CD4^{+}CD45RO^{+}CCR7^{+}CCR6^{+}CCR4^{+}$ |
| Central memory Th17.1 T cells | $CD4^{+}CD45RO^{+}CCR7^{+}CCR6^{+}CXCR3^{+}$ |
| Central memory gut-homing T cells | $CD4^{+}CD45RO^{+}CCR7^{+}\alpha4^{+}\beta7^{+}CCR9^{+}/CD8^{+}CD45RO^{+}CCR7^{+}\alpha4^{+}\beta7^{+}CCR9^{+}$ |
| Effector memory Th1 T cells | $CD4^{+}CD45RO^{+}CCR7^{-}CCR6^{-}CXCR3^{+}$ |
| Effector memory Th2 T cells | $CD4^{+}CD45RO^{+}CCR7^{-}CCR6^{-}CCR4^{+}$ |

| | |
|---|---|
| Effector memory Th17 T cells | $CD4^{+}CD45RO^{+}CCR7^{-}CCR6^{+}CCR4^{+}$ |
| Effector memory Th17.1 T cells | $CD4^{+}CD45RO^{+}CCR7^{-}CCR6^{+}CXCR3^{+}$ |
| Effector memory gut-homing T cells | $CD4^{+}CD45RO^{+}CCR7^{-}\alpha4^{+}\beta7^{+}CCR9^{+}$/$CD8^{+}CD45RO^{+}CCR7^{-}\alpha4^{+}\beta7^{+}CCR9^{+}$ |

Supplementary Table 3: Duodenal T cell populations in FD patients with and without concomitant IBS (%)

| | **Control (mean±SD)** | **FD IBS -ve (mean±SD)** | **FD IBS +ve (mean±SD)** | **Control vs. FD-IBS (*p* value)** | **Control vs. FD+IBS (*p* value)** | **FD-IBS vs. FD+IBS (*p* value)** |
|---|---|---|---|---|---|---|
| | n=23 | n=34 | n=15 | | | |
| CD3+ | 39.30±10.69 | 40.56±14.20 | 45.29±15.14 | 0.729 | 0.183 | 0.259 |
| CD4+ | 32.57±11.12 | 28.41±14.89 | 29.73±11.45 | 0.261 | 0.543 | 0.769 |
| CD8+ | 25.67±16.05 | 18.81±11.42 | 17.83±9.64 | 0.064 | 0.096 | 0.826 |
| CD4+ effector | 2.06±2.17 | 3.68±2.76 | 4.52±3.28 | 0.024* | 0.010* | 0.437 |
| CD4+ effector gut-homing | 5.04±5.46 | 7.39±6.15 | 6.97±4.92 | 0.138 | 0.202 | 0.944 |
| CD4+ effector Th1 | 7.90±13.22 | 1.87±4.15 | 1.28±2.62 | 0.165 | 0.251 | 0.994 |
| CD4+ effector Th2 | 13.03±16.11 | 19.03±16.80 | 21.67±12.42 | 0.098 | 0.035* | 0.417 |
| CD4+ effector Th17 | 31.74±24.72 | 46.33±24.69 | 43.84±22.19 | 0.029* | 0.137 | 0.741 |
| CD4+ effector Th17.1 | 14.17±25.75 | 3.78±133.67 | 0.50±0.98 | 0.633 | 0.365 | 0.578 |
| CD4+ naïve | 1.18±1.08 | 1.63±1.43 | 2.29±2.51 | 0.201 | 0.076 | 0.425 |
| CD8+ effector | 2.15±3.95 | 1.30±4.19 | 0.79±0.61 | 0.026* | 0.204 | 0.523 |
| CD8+ effector gut-homing | 3.23±5.79 | 1.70±2.69 | 1.27±2.44 | 0.547 | 0.422 | 0.733 |
| CD8+ naïve | 1.11±1.00 | 1.16±0.99 | 1.20±0.67 | 0.689 | 0.291 | 0.434 |
| CD4+ central memory | 3.35±2.68 | 2.37±1.66 | 2.229±1.55 | 0.083 | 0.132 | 0.909 |
| CD4+ central memory gut-homing | 19.19±10.08 | 17.74±11.14 | 20.72±12.92 | 0.641 | 0.686 | 0.398 |
| CD4+ central memory Th1 | 17.14±19.76 | 8.12±10.13 | 5.76±6.85 | 0.162 | 0.119 | 0.659 |
| CD4+ central memory Th2 | 23.75±18.97 | 37.20±17.73 | 38.23±17.72 | 0.014* | 0.038* | 0.941 |
| CD4+ central memory Th17 | 14.54±7.76 | 17.81±9.96 | 14.04±7.40 | 0.182 | 0.867 | 0.174 |

| | | | | | | |
|---|---|---|---|---|---|---|
| CD4+ central memory Th17.1 | 11.35±8.98 | 7.86±7.79 | 12.80±15.16 | 0.091 | 0.615 | 0.358 |
| CD4+ effector memory | 3.49±4.91 | 1.09±1.47 | 1.20±1.35 | 0.134 | 0.384 | 0.724 |
| CD4+ effector memory gut-homing | 9.08±8.24 | 3.97±4.49 | 5.29±4.93 | 0.015* | 0.248 | 0.378 |
| CD4+ effector memory Th1 | 16.14±15.65 | 10.41±8.96 | 6.58±6.30 | 0.510 | 0.100 | 0.232 |
| CD4+ effector memory Th2 | 9.80±10.50 | 17.85±11.50 | 26.45±17.76 | 0.023* | 0.000*** | 0.034* |
| CD4+ effector memory Th17 | 11.95±8.41 | 18.52±15.62 | 18.28±16.21 | 0.090 | 0.180 | 0.956 |
| CD4+ effector memory Th17.1 | 10.65±8.38 | 9.06±7.86 | 8.84±6.73 | 0.465 | 0.491 | 0.925 |
| CD8+ central memory | 2.78±1.59 | 3.10±2.38 | 3.03±2.00 | 0.577 | 0.718 | 0.916 |
| CD8+ central memory gut-homing | 16.17±16.45 | 5.21±5.99 | 3.16±3.07 | 0.005** | 0.003** | 0.468 |
| CD8+ effector memory | 5.70±8.79 | 2.17±4.74 | 0.70±0.68 | 0.449 | 0.282 | 0.618 |
| CD8+ effector memory gut-homing | 2.56±3.77 | 2.41±3.60 | 0.33±0.96 | 0.830 | 0.043* | 0.046* |

Supplementary Table 4: Peripheral T cell populations in FD patients with and without concomitant IBS (%)

| | **Control (mean±SD)** | **FD IBS -ve (mean±SD)** | **FD IBS +ve (mean±SD)** | **Control vs. FD-IBS (*p* value)** | **Control vs. FD+IBS (*p* value)** | **FD-IBS vs. FD+IBS (*p* value)** |
|---|---|---|---|---|---|---|
| | n=37 | n=33 | n=28 | | | |
| CD3+ | 41.13±13.70 | 48.44±26.06 | 47.31±10.87 | 0.101 | 0.184 | 0.811 |
| CD4+ | 45.63±18.70 | 40.51±17.89 | 47.23±13.26 | 0.220 | 0.708 | 0.133 |
| CD4+ gut-homing | 0.34±0.45 | 0.97±1.43 | 0.45±0.48 | 0.338 | 0.423 | 0.895 |
| CD8+ | 23.21±15.15 | 18.25±14.53 | 22.99±14.96 | 0.311 | 0.880 | 0.283 |
| CD8+ gut-homing | 0.364±0.267 | 0.573±0.60 | 0.55±0.42 | 0.363 | 0.152 | 0.621 |
| CD4+ effector | 1.59±2.07 | 1.28±1.73 | 1.62±1.49 | 0.466 | 0.298 | 0.091 |
| CD4+ effector gut-homing | 2.07±2.37 | 2.57±4.21 | 3.52±3.70 | 0.599 | 0.059 | 0.020* |
| CD4+ effector Th1 | 24.13±24.16 | 9.71±12.10 | 17.85±14.84 | 0.007** | 0.774 | 0.026* |
| CD4+ effector Th2 | 16.20±12.34 | 20.35±16.80 | 14.11±8.57 | 0.401 | 0.637 | 0.217 |
| CD4+ effector Th17 | 4.93±6.59 | 9.14±10.90 | 2.97±2.83 | 0.229 | 0.688 | 0.131 |
| CD4+ effector Th17.1 | 1.29±1.23 | 1.24±1.45 | 0.99±1.15 | 0.561 | 0.300 | 0.637 |
| CD4+ naïve | 15.35±12.94 | 17.26±13.90 | 18.00±13.90 | 0.557 | 0.438 | 0.834 |
| CD8+ effector | 6.07±6.24 | 2.10±3.25 | 3.28±3.00 | 0.003** | 0.400 | 0.048* |
| CD8+ effector gut-homing | 2.21±1.96 | 1.46±2.17 | 2.74±2.08 | 0.051 | 0.357 | 0.007** |
| CD8+ naïve | 5.06±4.24 | 7.95±7.40 | 7.34±7.80 | 0.161 | 0.319 | 0.731 |
| CD4+ central memory | 9.40±6.68 | 10.48±9.91 | 13.78±9.68 | 0.613 | 0.050 | 0.149 |
| CD4+ central memory gut-homing | 1.61±1.48 | 1.49±1.71 | 1.85±2.21 | 0.500 | 0.494 | 0.194 |
| CD4+ central memory Th1 | 9.63±6.44 | 15.17±11.12 | 15.80±11.36 | 0.057 | 0.029* | 0.721 |

| | | | | | | |
|---|---|---|---|---|---|---|
| CD4+ central memory Th2 | 11.79±7.12 | 16.35±13.34 | 11.64±7.78 | 0.349 | 0.731 | 0.224 |
| CD4+ central memory Th17 | 31.90±19.01 | 16.70±17.09 | 25.19±20.08 | 0.005** | 0.244 | 0.146 |
| CD4+ central memory Th17.1 | 19.75±15.10 | 7.01±11.80 | 17.92±16.65 | 0.002** | 0.748 | 0.011* |
| CD4+ effector memory | 4.13±4.06 | 2.03±2.57 | 4.11±3.65 | 0.043* | 0.872 | 0.035* |
| CD4+ effector memory gut-homing | 0.294±0.325 | 0.343±0.56 | 0.50±0.630 | 0.414 | 0.400 | 0.112 |
| CD4+ effector memory Th1 | 27.84±18.22 | 19.25±15.26 | 30.35±14.85 | 0.031* | 0.541 | 0.009** |
| CD4+ effector memory Th2 | 12.71±10.86 | 8.05±9.85 | 11.23±8.65 | 0.041* | 0.774 | 0.104 |
| CD4+ effector memory Th17 | 4.70±3.87 | 2.87±3.83 | 5.22±5.01 | 0.062 | 0.807 | 0.047* |
| CD4+ effector memory Th17.1 | 6.38±5.91 | 9.63±13.05 | 7.62±7.12 | 0.999 | 0.571 | 0.581 |
| CD8+ central memory | 2.20±2.08 | 2.29±2.31 | 2.46±2.17 | 0.995 | 0.679 | 0.700 |
| CD8+ central memory gut-homing | 4.11±3.19 | 3.42±3.87 | 4.31±3.48 | 0.216 | 0.942 | 0.218 |
| CD8+ effector memory | 1.67±1.54 | 0.963±1.23 | 1.99±1.99 | 0.108 | 0.678 | 0.055 |
| CD8+ effector memory gut-homing | 2.22±2.03 | 1.81±2.53 | 2.46±2.50 | 0.174 | 0.876 | 0.153 |

Supplementary Table 5: Duodenal T cell populations in FD patients and controls taking proton pump inhibitors (%)

| | **Control (mean±SD)** | **FD PPI -ve (mean±SD)** | **FD PPI +ve (mean±SD)** | **Control vs. FD -PPI (*p* value)** | **Control vs. FD +PPI (*p* value)** | **FD -PPI vs. FD +PPI (*p* value)** |
|---|---|---|---|---|---|---|
| | n=24 | n=21 | n=19 | | | |
| CD3+ | 38.90±10.64 | 43.16±14.64 | 43.74±13.41 | 0.273 | 0.226 | 0.888 |
| CD4+ | 32.46±10.88 | 27.84±13.09 | 29.87±16.38 | 0.281 | 0.536 | 0.657 |
| CD8+ | 25.83±15.72 | 17.01±10.21 | 20.39±12.49 | 0.042* | 0.200 | 0.458 |
| CD4+ effector | 1.99±2.15 | 4.17±3.08 | 4.19±3.12 | 0.011* | 0.013 | 0.973 |
| CD4+ effector gut-homing | 5.44±5.67 | 7.32±5.24 | 6.39±6.52 | 0.288 | 0.601 | 0.615 |
| CD4+ effector Th1 | 7.90±13.22 | 0.87±2.15 | 2.54±4.42 | 0.043* | 0.769 | 0.098 |
| CD4+ effector Th2 | 13.08±15.75 | 24.78±16.68 | 16.58±14.15 | 0.010* | 0.300 | 0.159 |
| CD4+ effector Th17 | 31.02±24.43 | 44.33±21.84 | 48.98±27.27 | 0.074 | 0.020* | 0.551 |
| CD4+ effector Th17.1 | 14.18±25.18 | 0.51±1.13 | 0.68±1.54 | 0.140 | 0.428 | 0.539 |
| CD4+ naïve | 1.15±1.07 | 1.75±1.31 | 2.68±2.82 | 0.091 | 0.29* | 0.589 |
| CD8+ effector | 2.20±3.93 | 0.63±0.44 | 0.67±0.55 | 0.018* | 0.028* | 0.920 |
| CD8+ effector gut-homing | 3.39±5.72 | 1.82±3.55 | 1.56±2.39 | 0.374 | 0.476 | 0.876 |
| CD8+ naïve | 1.10±0.98 | 1.23±0.89 | 1.47±1.30 | 0.479 | 0.251 | 0.650 |
| CD4+ central memory | 3.31±2.62 | 2.18±1.66 | 3.41±3.80 | 0.182 | 0.898 | 0.259 |
| CD4+ central memory gut-homing | 18.80±10.03 | 21.32±14.35 | 17.90±11.89 | 0.494 | 0.811 | 0.377 |
| CD4+ central memory Th1 | 17.35±19.35 | 4.63±5.31 | 11.29±13.68 | 0.039 | 0.464 | 0.211 |
| CD4+ central memory Th2 | 23.19±18.75 | 39.73±17.62 | 34.96±19.32 | 0.008** | 0.068 | 0.465 |
| CD4+ central memory Th17 | 14.97±7.86 | 19.94±8.49 | 14.03±9.32 | 0.050 | 0.905 | 0.047* |

| | | | | | | |
|---|---|---|---|---|---|---|
| CD4+ central memory Th17.1 | 11.12±8.84 | 8.52±9.34 | 8.30±7.22 | 0.167 | 0.318 | 0.738 |
| CD4+ effector memory | 3.39±4.82 | 0.59±0.48 | 1.71±2.05 | 0.011* | 0.796 | 0.034* |
| CD4+ effector memory gut-homing | 8.90±8.11 | 5.42±5.82 | 3.31±5.54 | 0.074 | 0.005** | 0.300 |
| CD4+ effector memory Th1 | 16.61±15.47 | 8.94±9.35 | 7.75±5.52 | 0.161 | 0.294 | 0.965 |
| CD4+ effector memory Th2 | 9.77±10.27 | 19.90±15.06 | 22.98±14.44 | 0.014* | 0.001** | 0.395 |
| CD4+ effector memory Th17 | 13.36±10.65 | 23.22±18.25 | 13.73±12.08 | 0.080 | 0.997 | 0.095 |
| CD4+ effector memory Th17.1 | 10.18±8.49 | 9.71±7.68 | 9.32±8.25 | 0.868 | 0.716 | 0.847 |
| CD8+ central memory | 2.80±1.55 | 2.92±2.58 | 3.42±2.07 | 0.850 | 0.352 | 0.462 |
| CD8+ central memory gut-homing | 16.02±16.11 | 3.55±3.88 | 4.90±6.32 | 0.001** | 0.007** | 0.697 |
| CD8+ effector memory | 6.28±9.06 | 0.48±0.57 | 2.39±4.66 | 0.032* | 0.730 | 0.100 |
| CD8+ effector memory gut-homing | 2.67±3.73 | 0.87±1.65 | 1.76±2.92 | 0.076 | 0.334 | 0.462 |

Supplementary Table 6: Peripheral T cell populations in FD patients and controls taking proton pump inhibitors (%)

| | **Control (mean±SD)** | **FD PPI -ve (mean±SD)** | **FD PPI +ve (mean±SD)** | **Control vs. FD -PPI (*p* value)** | **Control vs. FD +PPI (*p* value)** | **FD -PPI vs. FD +PPI (*p* value)** |
|---|---|---|---|---|---|---|
| | n=37 | n=37 | n=16 | | | |
| CD3+ | 41.13±13.70 | 48.64±18.36 | 49.90±26.42 | 0.194 | 0.023* | 0.205 |
| CD4+ | 45.63±18.70 | 46.16±16.19 | 39.24±11.72 | 0.891 | 0.215 | 0.181 |
| CD4+ gut-homing | 0.28±0.27 | 0.79±1.16 | 0.38±0.57 | 0.072 | 0.730 | 0.083 |
| CD8+ | 23.21±15.15 | 23.18±14.85 | 19.00±14.82 | 0.789 | 0.506 | 0.388 |
| CD8+ gut-homing | 0.34±0.27 | 0.57±0.50 | 0.52±0.38 | 0.159 | 0.260 | 0.970 |
| CD4+ effector | 1.59±2.07 | 1.47±1.72 | 1.16±1.17 | 0.843 | 0.603 | 0.501 |
| CD4+ effector gut-homing | 2.07±2.37 | 3.02±3.39 | 3.37±5.25 | 0.181 | 0.896 | 0.371 |
| CD4+ effector Th1 | 24.13±24.16 | 14.11±11.65 | 10.40±14.13 | 0.248 | 0.026* | 0.190 |
| CD4+ effector Th2 | 16.20±12.34 | 17.93±14.38 | 17.92±13.60 | 0.766 | 0.553 | 0.720 |
| CD4+ effector Th17 | 4.93±6.59 | 3.76±4.45 | 12.42±13.00 | 0.792 | 0.060 | 0.037* |
| CD4+ effector Th17.1 | 1.29±1.23 | 1.23±1.30 | 1.07±1.39 | 0.743 | 0.374 | 0.525 |
| CD4+ naïve | 15.35±12.94 | 19.60±13.46 | 17.61±15.88 | 0.186 | 0.584 | 0.628 |
| CD8+ effector | 6.08±6.24 | 3.31±3.76 | 2.54±3.76 | 0.129 | 0.035* | 0.359 |
| CD8+ effector gut-homing | 2.21±1.96 | 2.30±2.19 | 1.58±2.59 | 0.972 | 0.128 | 0.136 |
| CD8+ naïve | 5.06±4.24 | 9.15±7.95 | 6.40±7.21 | 0.041* | 0.745 | 0.220 |
| CD4+ central memory | 9.40±6.68 | 12.16±8.60 | 16.51±14.81 | 0.196 | 0.123 | 0.592 |
| CD4+ central memory gut-homing | 1.61±1.48 | 1.65±2.07 | 1.76±2.04 | 0.939 | 0.711 | 0.668 |
| CD4+ central memory Th1 | 9.63±6.44 | 16.78±11.78 | 11.02±6.36 | 0.001** | 0.619 | 0.041* |

| | | | | | | |
|---|---|---|---|---|---|---|
| CD4+ central memory Th2 | 11.79±7.12 | 11.42±6.90 | 15.88±15.24 | 0.860 | 0.135 | 0.103 |
| CD4+ central memory Th17 | 31.90±19.01 | 24.67±19.70 | 13.47±16.29 | 0.195 | 0.005** | 0.073 |
| CD4+ central memory Th17.1 | 19,75±15.10 | 17.96±16.01 | 3.39±8.49 | 0.857 | 0.000*** | 0.001** |
| CD4+ effector memory | 4.13±4.06 | 3.72±3.52 | 1.66±2.11 | 0.957 | 0.036* | 0.039* |
| CD4+ effector memory gut-homing | 0.29±0.32 | 0.49±0.63 | 0.31±0.57 | 0.443 | 0.333 | 0.121 |
| CD4+ effector memory Th1 | 27.84±18.22 | 28.29±14.23 | 15.59±15.31 | 0.906 | 0.013* | 0.010* |
| CD4+ effector memory Th2 | 12.71±10.86 | 10.93±8.04 | 9.29±12.82 | 0.457 | 0.266 | 0.596 |
| CD4+ effector memory Th17 | 4.70±3.87 | 5.01±4.74 | 2.12±3.69 | 0.870 | 0.026* | 0.018* |
| CD4+ effector memory Th17.1 | 6.38±5.91 | 10.19±11.47 | 8.02±10.69 | 0.280 | 0.759 | 0.249 |
| CD8+ central memory | 2.20±2.08 | 2.42±2.11 | 2.98±2.60 | 0.693 | 0.303 | 0.469 |
| CD8+ central memory gut-homing | 4.11±3.19 | 3.73±3.58 | 4.07±3.85 | 0.640 | 0.965 | 0.747 |
| CD8+ effector memory | 1.67±1.54 | 1.73±1.88 | 0.88±1.05 | 0.901 | 0.193 | 0.225 |
| CD8+ effector memory gut-homing | 2.22±2.03 | 1.95±2.13 | 3.17±3.41 | 0.509 | 0.710 | 0.380 |

Supplementary Table 7: Duodenal T cell populations in FD patients and controls analysed by sex (%)

| | **Control (Female) (mean±SD)** | **Control (Male) (mean±SD)** | **FD (Female) (mean±SD)** | **FD (Male) (mean±SD)** | **Control female vs. Control male (*p* value)** | **Control female vs. FD female (*p* value)** | **Control female vs. FD male (*p* value)** | **Control male vs. FD female (*p* value)** | **Control male vs. FD male (*p* value)** | **FD female vs. FD male (*p* value)** |
|---|---|---|---|---|---|---|---|---|---|---|
| | n=11 | n=12 | n=37 | n=11 | | | | | | |
| CD3+ | 44.99±9.14 | 36.24±6.25 | 42.91±15.45 | 39.04±11.73 | 0.120 | 0.643 | 0.287 | 0.140 | 0.616 | 0.390 |
| CD4+ | 30.85±11.15 | 34.13±111.34 | 29.97±13.98 | 25.74±13.54 | 0.548 | 0.848 | 0.360 | 0.354 | 0.127 | 0.361 |
| CD8+ | 22.76±15.63 | 28.34±16.64 | 19.25±12.41 | 16.55±3.99 | 0.572 | 0.407 | 0.406 | 0.122 | 0.158 | 0.861 |
| CD4+ effector | 2.52±2.46 | 1.20±0.99 | 4.27±3.57 | 3.73±2.21 | 0.225 | 0.152 | 0.223 | 0.002** | 0.013* | 0.949 |
| CD4+ effector gut-homing | 3.93±4.21 | 5.96±6.34 | 6.80±5.64 | 7.89±5.77 | 0.402 | 0.156 | 0.111 | 0.653 | 0.412 | 0.573 |
| CD4+ effector Th1 | 4.31±10.51 | 7.52±9.67 | 1.69±3.72 | 0.70±1.48 | 0.293 | 0.834 | 0.731 | 0.121 | 0.161 | 0.825 |
| CD4+ effector Th2 | 11.66±9.95 | 14.28±20.62 | 21.53±15.47 | 14.03±15.68 | 0.692 | 0.073 | 0.726 | 0.172 | 0.969 | 0.171 |
| CD4+ effector Th17 | 44.07±28.25 | 20.44±14.32 | 45.15±24.39 | 46.21±23.63 | 0.019* | 0.894 | 0.832 | 0.002** | 0.011* | 0.896 |
| CD4+ effector Th17.1 | 0.51±0.79 | 23.27±31.57 | 1.49±3.52 | 0.36±0.33 | 0.185 | 0.700 | 0.537 | 0.202 | 0.497 | 0.705 |
| CD4+ naïve | 1.57±1.30 | 0.63±0.35 | 1.92±2.02 | 2.07±2.37 | 0.074 | 0.777 | 0.742 | 0.0126* | 0.038* | 0.900 |
| CD8+ effector | 0.96±0.84 | 4.89±7.41 | 1.29±4.02 | 0.73±0.38 | 0.143 | 0.293 | 0.908 | 0.004** | 0.114 | 0.363 |
| CD8+ effector gut-homing | 0.91±2.17 | 2.46±3.71 | 1.79±3.04 | 1.85±2.72 | 0.120 | 0.269 | 0.432 | 0.410 | 0.431 | 0.880 |
| CD8+ naïve | 1.49±1.32 | 0.75±0.36 | 1.49±2.06 | 1.07±0.62 | 0.180 | 0.780 | 0.820 | 0.162 | 0.282 | 0.992 |
| CD4+ central memory | 2.40±2.01 | 4.14±2.98 | 2.62±2.96 | 2.60±1.50 | 0.172 | 0.987 | 0.591 | 0.083 | 0.402 | 0.506 |
| CD4+ central memory gut-homing | 17.07±9.70 | 20.95±10.47 | 19.96±13.22 | 18.49±12.14 | 0.442 | 0.792 | 0.987 | 0.479 | 0.447 | 0.811 |

| | | | | | | | | | | |
|---|---|---|---|---|---|---|---|---|---|---|
| CD4+ central memory Th1 | 47.10±7.41 | 25.93±21.89 | 8.15±9.71 | 3.25±1.95 | 0.004** | 0.344 | 0.663 | 0.006** | 0.014* | 0.689 |
| CD4+ central memory Th2 | 27.20±20.82 | 20.59±17.39 | 36.92±18.32 | 38.90±16.14 | 0.390 | 0.126 | 0.138 | 0.009** | 0.019* | 0.753 |
| CD4+ central memory Th17 | 15.83±8.99 | 11.66±2.79 | 16.56±10.20 | 17.73±3.54 | 0.266 | 0.812 | 0.619 | 0.099 | 0.107 | 0.701 |
| CD4+ central memory Th17.1 | 5.89±5.39 | 17.54±12.00 | 9.16±10.64 | 8.81±7.30 | 0.007** | 0.547 | 0.353 | 0.0047** | 0.067 | 0.579 |
| CD4+ effector memory | 0.78±1.02 | 4.77±5.14 | 1.08±1.33 | 1.13±0.79 | 0.004** | 0.383 | 0.209 | 0.006** | 0.117 | 0.486 |
| CD4+ effector memory gut-homing | 10.97±8.86 | 7.36±6.37 | 4.42±5.05 | 3.89±3.06 | 0.157 | 0.003** | 0.008** | 0.150 | 0.174 | 0.799 |
| CD4+ effector memory Th1 | 12.37±14.90 | 19.60±16.15 | 9.88±10.42 | 8.91±2.08 | 0.197 | 0.900 | 0.662 | 0.080 | 0.417 | 0.512 |
| CD4+ effector memory Th2 | 7.15±8.33 | 12.23±11.99 | 20.10±15.98 | 20.80±5.47 | 0.300 | 0.005** | 0.002** | 0.115 | 0.035* | 0.301 |
| CD4+ effector memory Th17 | 10.84±10.75 | 13.06±5.52 | 18.18±15.46 | 19.11±17.62 | 0.516 | 0.165 | 0.244 | 0.562 | 0.607 | 0.954 |
| CD4+ effector memory Th17.1 | 10.49±9.11 | 10.80±8.04 | 8.87±7.95 | 9.07±6.23 | 0.929 | 0.554 | 0.675 | 0.483 | 0.611 | 0.942 |
| CD8+ central memory | 2.57±1.77 | 2.95±1.47 | 2.97±2.38 | 3.08±1.57 | 0.666 | 0.587 | 0.573 | 0.978 | 0.883 | 0.880 |
| CD8+ central memory gut-homing | 12.33±15.93 | 19.69±16.80 | 5.31±5.96 | 2.54±2.04 | 0.068 | 0.337 | 0.091 | 0.001** | 0.000*** | 0.255 |
| CD8+ effector memory | 0.98±1.80 | 7.63±8.66 | 2.25±4.96 | 1.29±1.35 | 0.013* | 0.294 | 0.139 | 0.039* | 0.319 | 0.430 |
| CD8+ effector memory gut-homing | 0.78±2.08 | 3.32±3.64 | 1.60±2.76 | 2.69±4.50 | 0.036* | 0.463 | 0.215 | 0.058 | 0.283 | 0.417 |

Supplementary Table 8: Peripheral T cell populations in FD patients and controls analysed by sex (%)

| | **Control (Female) (mean±SD)** | **Control (Male) (mean±SD)** | **FD (Female) (mean±SD)** | **FD (Male) (mean±SD)** | **Control female vs. Control male (*p* value)** | **Control female vs. FD female (*p* value)** | **Control female vs. FD male (*p* value)** | **Control male vs. FD female (*p* value)** | **Control male vs. FD male (*p* value)** | **FD female vs. FD male (*p* value)** |
|---|---|---|---|---|---|---|---|---|---|---|
| | n=19 | n=18 | n=42 | n=19 | | | | | | |
| CD3+ | 42.83±12.02 | 39.44±15.36 | 48.83±20.89 | 45.90±19.64 | 0.887 | 0.198 | 0.436 | 0.146 | 0.356 | 0.701 |
| CD4+ | 42.87±20.56 | 48.54±16.60 | 44.11±16.89 | 42.76±14.50 | 0.319 | 0.795 | 0.985 | 0.365 | 0.317 | 0.782 |
| CD4+ gut-homing | 0.28±0.33 | 0.28±0.21 | 0.70±0.96 | 0.53±1.05 | 0.645 | 0.144 | 0.793 | 0.372 | 0.844 | 0.260 |
| CD8+ | 17.96±12.36 | 28.76±16.15 | 17.35±13.81 | 28.08±14.71 | 0.033* | 0.932 | 0.035* | 0.018* | 0.970 | 0.020* |
| CD8+ gut-homing | 0.35±0.30 | 0.38±0.24 | 0.61±0.61 | 0.48±0.37 | 0.598 | 0.151 | 0.352 | 0.411 | 0.662 | 0.797 |
| CD4+ effector | 2.17±2.84 | 1.04±0.96 | 1.531±1.53 | 1.22±1.85 | 0.420 | 0.961 | 0.347 | 0.375 | 0.907 | 0.396 |
| CD4+ effector gut-homing | 1.43±1.90 | 2.33±2.16 | 3.93±5.89 | 1.87±2.36 | 0.186 | 0.070 | 0.582 | 0.820 | 0.433 | 0.243 |
| CD4+ effector Th1 | 26.44±27.82 | 18.19±13.97 | 12.18±11.68 | 16.23±18.01 | 0.930 | 0.193 | 0.409 | 0.176 | 0.373 | 0.740 |
| CD4+ effector Th2 | 12.80±9.32 | 17.82±12.66 | 17.96±13.25 | 15.11±10.86 | 0.254 | 0.190 | 0.753 | 0.975 | 0.409 | 0.347 |
| CD4+ effector Th17 | 4.95±7.32 | 3.71±3.225 | 5.33±8.11 | 8.60±10.02 | 0.580 | 0.708 | 0.088 | 0.779 | 0.263 | 0.102 |
| CD4+ effector Th17.1 | 0.72±0.783 | 1.87±1.35 | 1.02±1.35 | 1.34±1.44 | 0.009** | 0.580 | 0.216 | 0.013* | 0.169 | 0.364 |
| CD4+ naïve | 14.44±13.40 | 16.32±12.75 | 19.72±14.92 | 11.32±6.92 | 0.663 | 0.148 | 0.470 | 0.358 | 0.254 | 0.025* |
| CD8+ effector | 4.14±4.44 | 7.19±6.68 | 2.29±2.82 | 4.15±4.67 | 0.162 | 0.255 | 0.749 | 0.005** | 0.280 | 0.129 |
| CD8+ effector gut-homing | 2.17±2.18 | 2.25±1.76 | 2.37±3.32 | 2.13±1.90 | 0.720 | 0.704 | 0.907 | 0.429 | 0.808 | 0.606 |
| CD8+ naïve | 4.71±4.68 | 5.44±3.82 | 7.40±7.23 | 8.68±8.27 | 0.393 | 0.167 | 0.095 | 0.716 | 0.427 | 0.568 |
| CD4+ central memory | 9.35±6.96 | 9.47±6.55 | 13.18±11.56 | 11.32±8.91 | 0.960 | 0.390 | 0.470 | 0.375 | 0.451 | 0.992 |

| | | | | | | | | | | |
|---|---|---|---|---|---|---|---|---|---|---|
| CD4+ central memory gut-homing | 1.68±1.67 | 1.29±0.83 | 2.14±2.78 | 0.98±0.61 | 0.910 | 0.573 | 0.367 | 0.493 | 0.437 | 0.103 |
| CD4+ central memory Th1 | 9.79±7.43 | 9.46±5.50 | 17.44±11.78 | 9.656±5.52 | 0.914 | 0.004** | 0.964 | 0.003** | 0.964 | 0.003** |
| CD4+ central memory Th2 | 12.88±8.68 | 10.57±4.82 | 15.76±12.08 | 10.70±8.65 | 0.544 | 0.636 | 0.306 | 0.246 | 0.700 | 0.094 |
| CD4+ central memory Th17 | 30.26±19.23 | 33.63±19.17 | 16.46±17.09 | 29.75±19.78 | 0.580 | 0.008** | 0.932 | 0.001** | 0.524 | 0.011* |
| CD4+ central memory Th17.1 | 17.79±16.48 | 21.82±13.65 | 11.08±15.33 | 16.35±16.61 | 0.415 | 0.195 | 0.864 | 0.026* | 0.325 | 0.274 |
| CD4+ effector memory | 3.22±4.18 | 5.05±3.83 | 2.45±2.69 | 4.81±4.65 | 0.102 | 0.760 | 0.218 | 0.025* | 0.671 | 0.076 |
| CD4+ effector memory gut-homing | 0.25±0.32 | 0.34±0.33 | 0.63±1.39 | 0.28±0.33 | 0.366 | 0.560 | 0.803 | 0.631 | 0.518 | 0.779 |
| CD4+ effector memory Th1 | 28.83±19.00 | 26.80±17.85 | 24.81±15.77 | 23.33±16.75 | 0.717 | 0.394 | 0.321 | 0.679 | 0.536 | 0.753 |
| CD4+ effector memory Th2 | 11.72±10.95 | 13.76±10.98 | 9.31±8.98 | 10.05±10.46 | 0.537 | 0.389 | 0.615 | 0.119 | 0.270 | 0.795 |
| CD4+ effector memory Th17 | 3.90±3.86 | 5.59±3.81 | 3.37±3.37 | 5.35±5.56 | 0.204 | 0.686 | 0.509 | 0.062 | 0.541 | 0.243 |
| CD4+ effector memory Th17.1 | 7.52±6.98 | 5.09±4.27 | 10.05±11.65 | 4.24±3.81 | 0.436 | 0.855 | 0.233 | 0.282 | 0.696 | 0.118 |
| CD8+ central memory | 1.89±1.84 | 2.07±1.46 | 2.49±2.44 | 2.11±1.72 | 0.607 | 0.512 | 0.581 | 0.970 | 0.982 | 0.991 |
| CD8+ central memory gut-homing | 4.37±3.82 | 3.85±2.49 | 3.86±3.92 | 3.78±3.25 | 0.662 | 0.610 | 0.612 | 0.995 | 0.949 | 0.935 |
| CD8+ effector memory | 1.00±0.89 | 2.21±1.73 | 1.06±1.34 | 2.24±2.05 | 0.063 | 0.941 | 0.057 | 0.024* | 0.993 | 0.019* |
| CD8+ effector memory gut-homing | 1.83±1.99 | 2.66±2.06 | 2.16±2.70 | 2.02±2.13 | 0.277 | 0.980 | 0.860 | 0.200 | 0.360 | 0.818 |

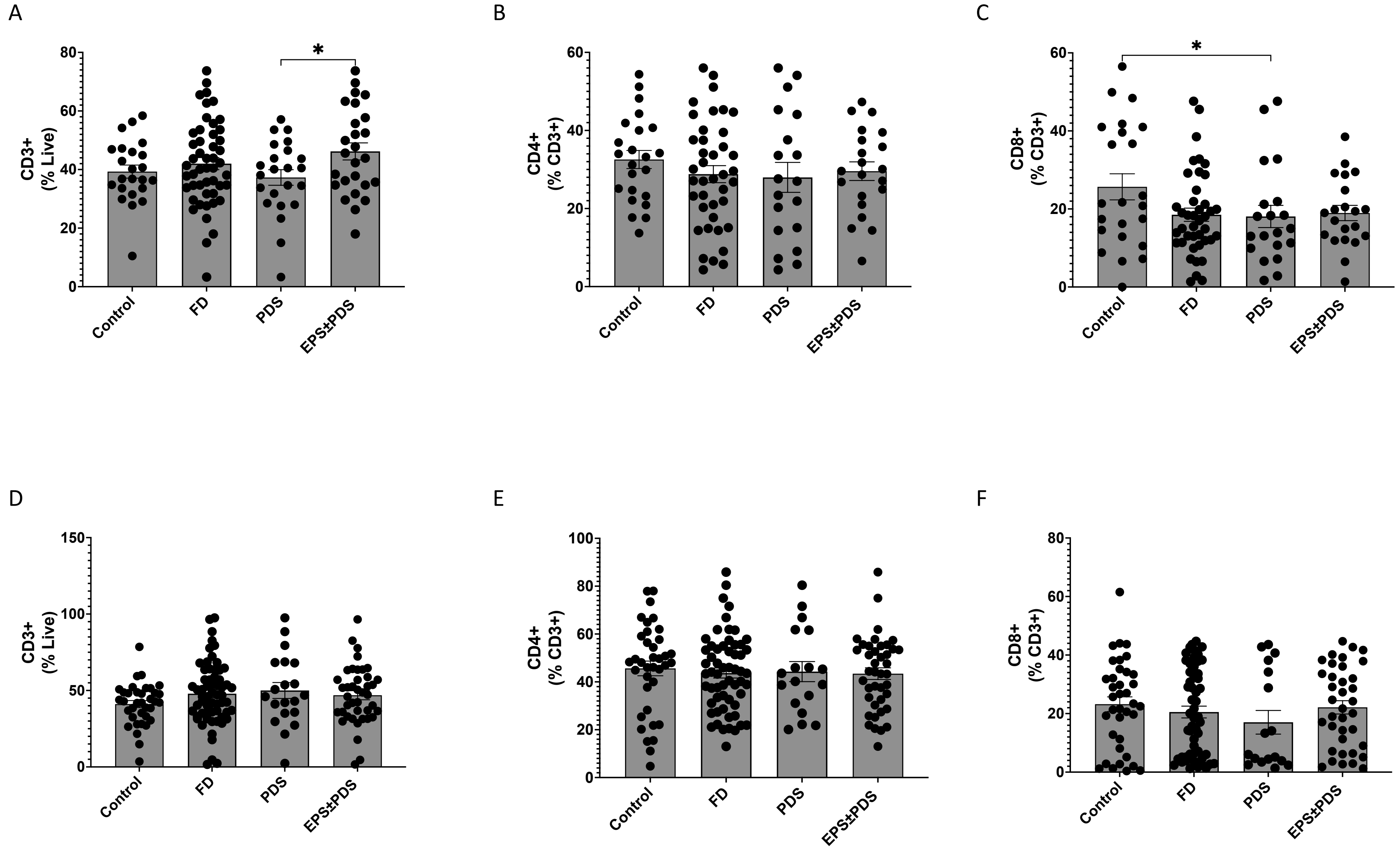
A
B
C
D
E
F
CD3+ (% Live)
CD4+ (% CD3+)
CD8+ (% CD3+)
*
Control
FD
PDS
EPS±PDS

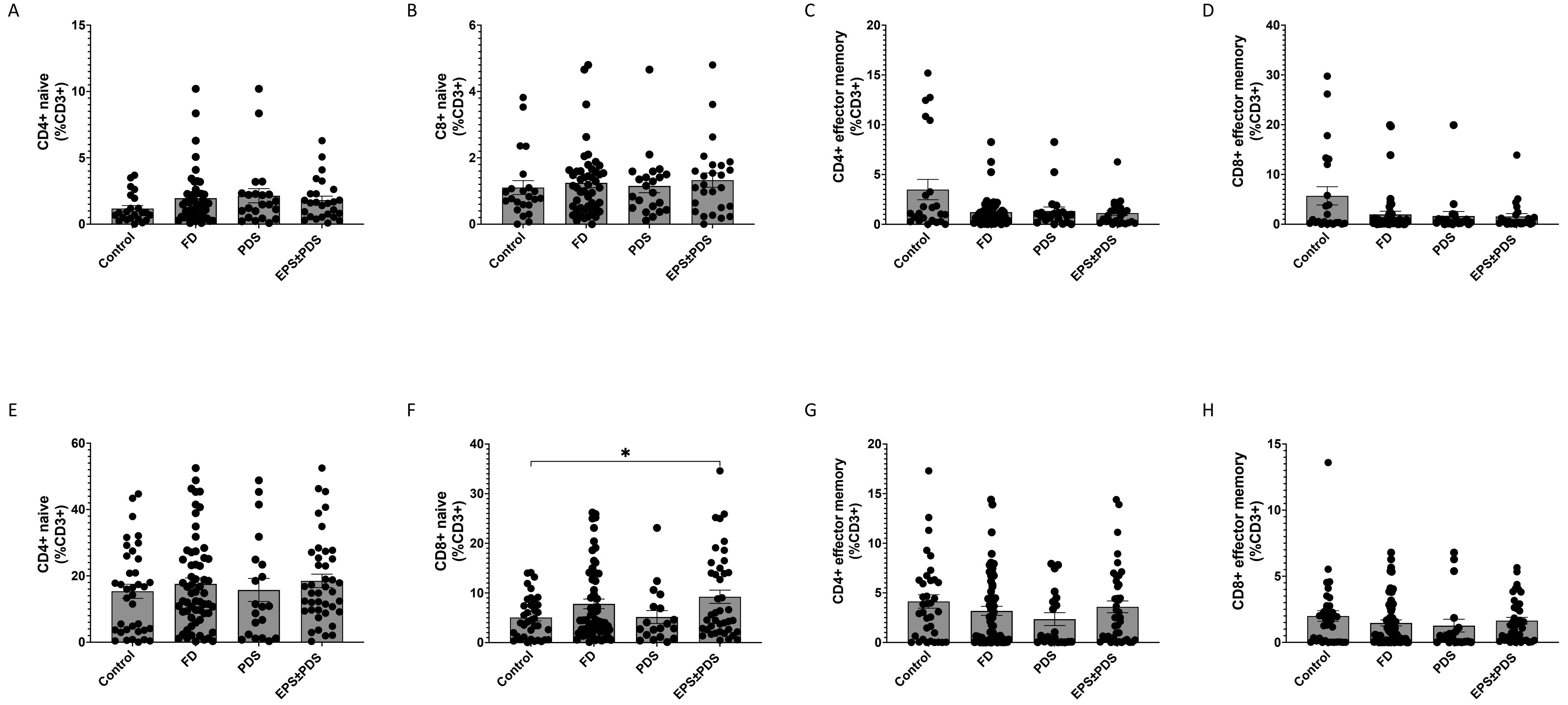
A
B
C
D
E
F
G
H
CD4+ naive (%CD3+)
C8+ naive (%CD3+)
CD4+ effector memory (%CD3+)
CD8+ effector memory (%CD3+)
CD4+ naive (%CD3+)
CD8+ naive (%CD3+)
CD4+ effector memory (%CD3+)
CD8+ effector memory (%CD3+)
Control
FD
PDS
EPS±PDS
*

Supplementary Figure 3

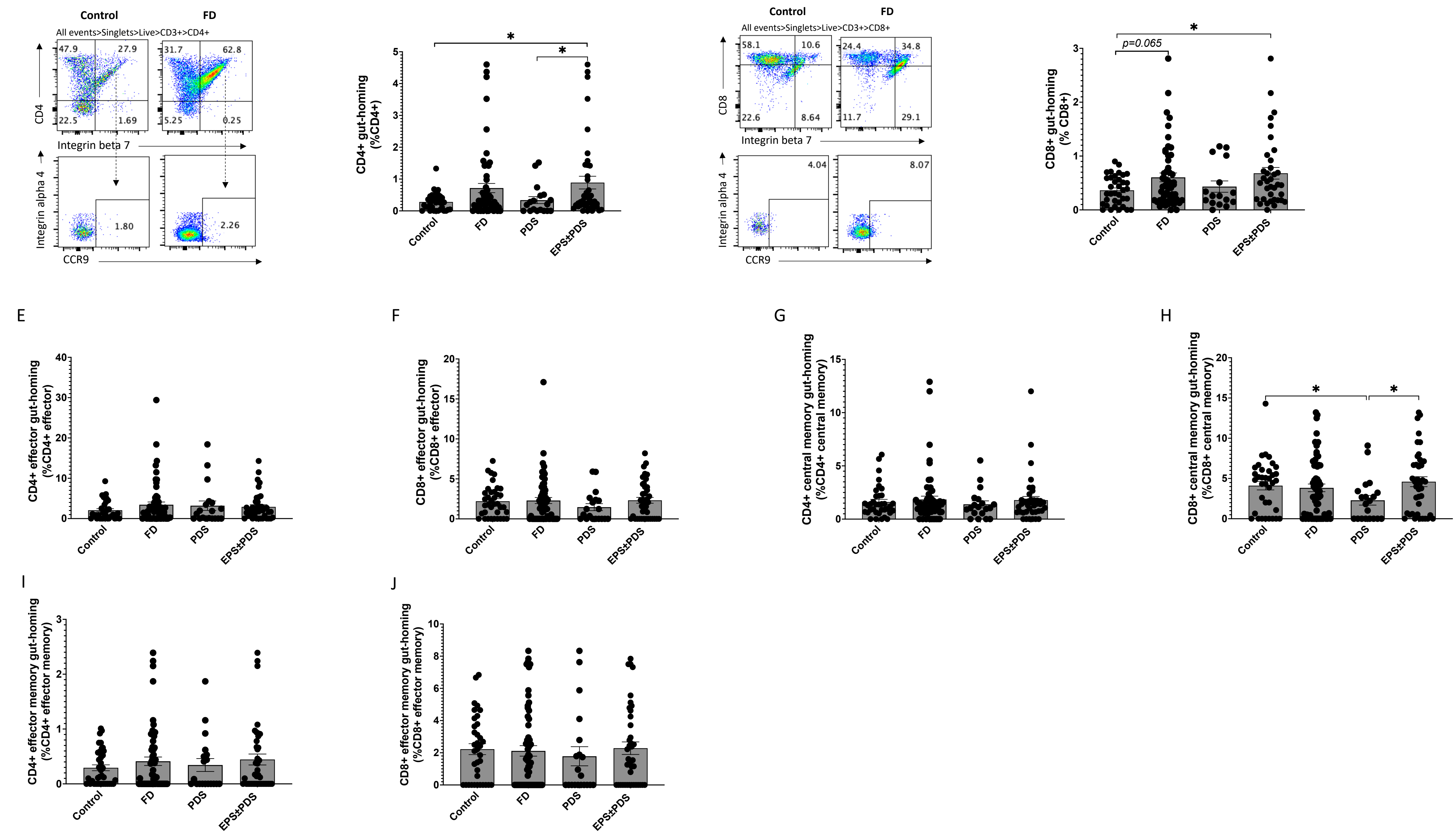

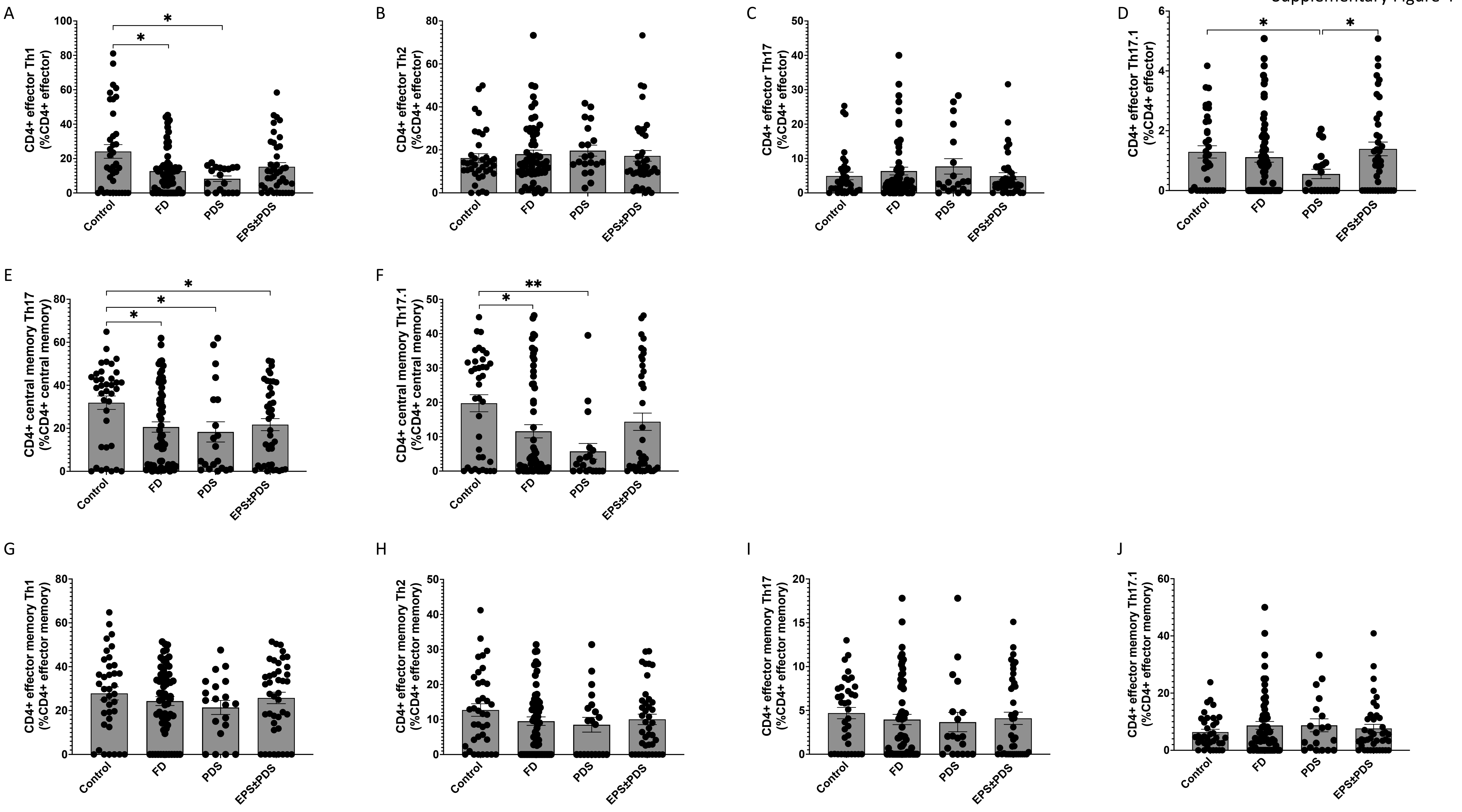
A
CD4+ effector Th1 (%CD4+ effector)
B
CD4+ effector Th2 (%CD4+ effector)
C
CD4+ effector Th17 (%CD4+ effector)
D
CD4+ effector Th17.1 (%CD4+ effector)
E
CD4+ central memory Th17 (%CD4+ central memory)
F
CD4+ central memory Th17.1 (%CD4+ central memory)
G
CD4+ effector memory Th1 (%CD4+ effector memory)
H
CD4+ effector memory Th2 (%CD4+ effector memory)
I
CD4+ effector memory Th17 (%CD4+ effector memory)
J
CD4+ effector memory Th17.1 (%CD4+ effector memory)
Control
FD
PDS
EPS±PDS
*
**
